# Decoding menstrual health across the lifespan: a scoping review of digital health tools in research

**DOI:** 10.1101/2025.09.24.25336575

**Authors:** Sarah C Johnson, Johanna O’Day, Emily Kraus, Scott Delp, Jennifer Hicks

## Abstract

Digital health tools provide longitudinal physiological and behavioural data that can address knowledge gaps in women’s health. This is particularly relevant for understanding hormone-driven physiological changes and symptoms, which impact health and performance across the lifespan. We conducted a scoping review of research using wearables or smartphone applications to identify insights about physiology, health behaviours, and symptoms throughout the menstrual cycle and menopausal transition. We identified 40 original articles. We summarise findings that reproduce lab-based results, giving confidence in the use of digital health tools for studying menstrual health, along with new insights gained. Given the importance of validation against gold standards, and the lack of a prior synthesis of wearable accuracy for women’s health applications, we next report accuracies of wearables that measure biometrics relevant to menstrual health. Finally, we discuss future research needs, including understanding physiological changes during perimenopause, and the role of health behaviours in symptom management.

## 1. Introduction

Women experience hormonal fluctuations throughout the menstrual cycle and across the lifespan. But there is a lack of knowledge about how to effectively manage associated symptoms and optimise health, athletic performance, and productivity. Between 74 and 90% of women report symptoms, both prior to and during menses, for example mood swings and abdominal bloating or dysmenorrhea (pain during the menstrual cycle)^1–3^, with 20 to 40% estimated to experience premenstrual syndrome (PMS)^2^. Furthermore 3 to 8% of women report severe and disabling premenstrual symptoms that meet the criteria for premenstrual dysphoric disorder (PMDD)^2,4^.

These symptoms associated with the menstrual cycle can affect quality of life. For example, in a study of over 32,000 women in the Netherlands, 81% reported impaired work performance in the past three months due to the menstrual cycle, and 14% reported absenteeism in the past six months^5^. Another study found 45% of surveyed women using the Flo app and living in the United States, reported absenteeism related to the menstrual cycle in the previous twelve months^6^, while in low and middle income countries there was pooled absenteeism of 15%^7^. In Japan, the estimated annual economic burden due to menstrual symptoms was estimated at 8.6 billion United States Dollars^1^. Across a range of sports, more than 50% of female athletes reported a negative impact of menstrual and premenstrual symptoms on training and competition^8–10^. Further, perimenopausal women have 40% higher odds of displaying depressive symptoms than premenopausal women^11^, and consistently show moderately impaired quality of life^12^. The economic burden due to menopausal transition (perimenopause) symptoms cost the United States more than $26 billion annually^13^.

The cyclical changes in hormones are well characterised throughout the menstrual cycle: during the follicular phase, beginning with menses, increasing concentrations of follicle stimulating hormone (FSH) stimulate ovarian follicles to secrete estrogen. Once a critical estrogen threshold is reached, a surge in luteinizing hormone (LH) and a smaller spike in FSH occurs, triggering ovulation. In the subsequent luteal phase, progesterone is secreted from the corpus luteum that releases the egg, LH and FSH concentrations decline, and estrogen concentration remains high. Subsequent decline of progesterone and estrogen triggers menses. The gold standard for ovulation confirmation is transvaginal ultrasound imaging of follicular release, with serum testing to confirm the LH surge and progesterone peak as the second preferred choice. However, LH testing alone (typically in urine) and/or tracking of basal body temperature increases at ovulation are commonly used as non-invasive measures.

The use of hormonal contraception, as well as the menopausal transition, alters endogenous hormonal profiles in different ways. Many contraceptives involve the exogenous administration of hormones. For example, combined oral contraceptives release synthetic estrogen and progesterone systemically, which suppresses endogenous ovarian hormone production^14^. The hormonal IUD secretes synthetic progestin which acts locally to cause endometrial thinning, inhibition of sperm motility, and cervical mucus thickening, but may also suppress endogenous hormones, and inhibit ovulation in some women^15^. During perimenopause, rising FSH concentrations reflect a declining number of ovarian follicles and responsiveness, resulting in pronounced estrogen fluctuations before permanent decline^16,17^.

Findings from lab studies have identified some connections between these hormonal changes in the menstrual cycle and physiological measures and symptoms. Lab studies have shown that progesterone consistently elevates core body temperature across various female cohorts^18,19^.

This effect, consistent with the observed high progesterone and higher core temperature in the luteal phase, appears to be reduced by estrogen^20^. During the luteal phase, heart rate also increases, potentially due to the influence of progesterone and estrogen on the autonomic nervous system or as a secondary effect of elevated body temperature^21^. Progesterone increases respiratory function by increasing carbon dioxide sensitivity, breathing rate, and potentially bronchodilation via smooth muscle relaxation, while evidence of estrogen’s interaction on this effect is contradictory^22,23^. Understanding links between symptoms and physiology is limited, but several lab-based studies suggest that women with severe premenstrual symptoms or PMDD often exhibit lower heart rate variability (HRV), indicating reduced parasympathetic activity, at different times in the menstrual cycle^24–27^. While these findings are valuable, our knowledge of the relationships between hormones, physiology, symptoms, and health behaviours is still limited.

Advances in digital health tools, including wearable technology and smartphone apps, provide a unique opportunity to bridge critical knowledge gaps in female health, wellbeing, and performance by enabling large-scale, continuous, and cost-effective collection of physiological and behavioural data. In-lab approaches, while valuable, have been limited by sample size and diversity, measurement frequency, and a lack of accompanying physical activity and sleep data. Wearables can continuously measure heart rate, HRV, respiratory rate, skin temperature, sleep metrics, and activity metrics. Independent or associated smartphone apps allow users to record menses dates, daily symptoms, mental state, as well as activity and behaviour details. Digital health tools also include apps with non-wearable devices, for instance, temporary smartphone camera add-ons can be used with an app to provide quantitative readings of hormonal urine test strips^28^.

Given the proliferation of digital health tools, there is a need to evaluate how they are being used in women’s health research. It is important to assess their accuracy, the discoveries they have enabled, and opportunities for future research. Past reviews of digital tools addressing women’s health have focused on identifying unmet needs in software design and hardware development^29–31^, accuracy of predicting ovulation and menses onset for contraception and conception purposes^30,32–35^, and the utility and implementation of technology tailored to monitor and diagnose conditions prevalent in women^36–38^. We found only one review that summarised findings from passive monitoring with commercial wearables, but this review focused solely on trends in thermoregulation relevant to the menstrual cycle^21^.

No existing reviews summarise findings from digital health tools that concurrently track female-specific, hormone-induced events such as the menstrual cycle or menopausal transition and report physiological, behavioural, and symptomatic outputs. Further, no prior review has systematically evaluated whether digital technology has led to significant new research insights. There is also a critical need to understand when women’s health researchers can trust digital health tools, which can be assessed by both direct validation compared to gold standard measures, as well as indirect validation to determine if tools reproduce known relationships from lab-based studies. While many studies and reviews have evaluated the accuracy of individual wearable sensors to predict characteristics related to the menstrual cycle (e.g. length, phase), none have characterised accuracy in the context of women’s health research or across multiple physiological biometrics and behaviours (i.e., resting heart rate, HRV, respiratory rate, skin temperature, and sleep).

We conducted a scoping review of studies that used digital health tools—including mobile applications, wearable devices, or integrated app-device systems—to characterise changes associated with female-specific, hormone-driven events such as the menstrual cycle and menopausal transition. Our review includes menstruating females, individuals using hormonal contraceptives, and those navigating the menopausal transition. The review excludes findings regarding pregnancy and postpartum. While we did not exclude studies that included women with gynecological conditions such as endometriosis or gynecological cancer, these conditions were not the focus of the review.

We first summarise the knowledge gained from digital health studies about the interplay between physiology, behaviours, symptoms, and hormonally driven processes. We discuss new insights gained, and compare findings from digital health studies to prior lab-based work. We then summarise the reported accuracy and resolution of digital health tools to assist researchers in making informed decisions about device selection and digital health study design. We close by discussing key unmet research needs in female health, wellbeing, and performance that could be addressed using digital health tools in order to inspire future research.

## 2. Results

### 2.1 Characteristics of included studies

In total, 40 studies were considered eligible for this scoping review based on the Preferred Reporting Items for Systematic Reviews and Meta-Analyses extension for Scoping Reviews (PRISMA) process (see Methods). Our search terms related to digital health tools (“digital health” OR “wearable technology” OR “wearable devices” OR “mobile health” OR “smartphone application” OR “app”) and menstrual health (“ovulation” OR “ovulatory cycle” OR “cycle tracking”), (“menstrual cycle” OR “menstruation” OR “period tracking”) identified 168 original research articles. We screened the abstracts of all 168 identified articles, screened the full text of 51 articles, and ultimately included 23 of the identified articles in our review. From the references of these identified articles and websites, we extracted an additional 43 papers and included 16 additional articles to bring the total from 23 to 39. Our search for digital health studies of perimenopause and menopause (“menopaus*” OR “perimenopaus*”) resulted in 15 articles for screening, and only four for inclusion, of which only one was unique to the menopause search, bringing the eligible articles total from 39 to 40. Study cohorts ranged in size from 10 to 19,000,000 female participants, predominantly aged 18 to 55, but in a few studies participants were as young as 13 or as old as 90. We summarised reported non-biometric menstrual characteristics (Table 1; 17 relevant studies) and biometric and behavioural insights (Table 2; 29 relevant studies) across the menstrual cycle.

**Table 1:**
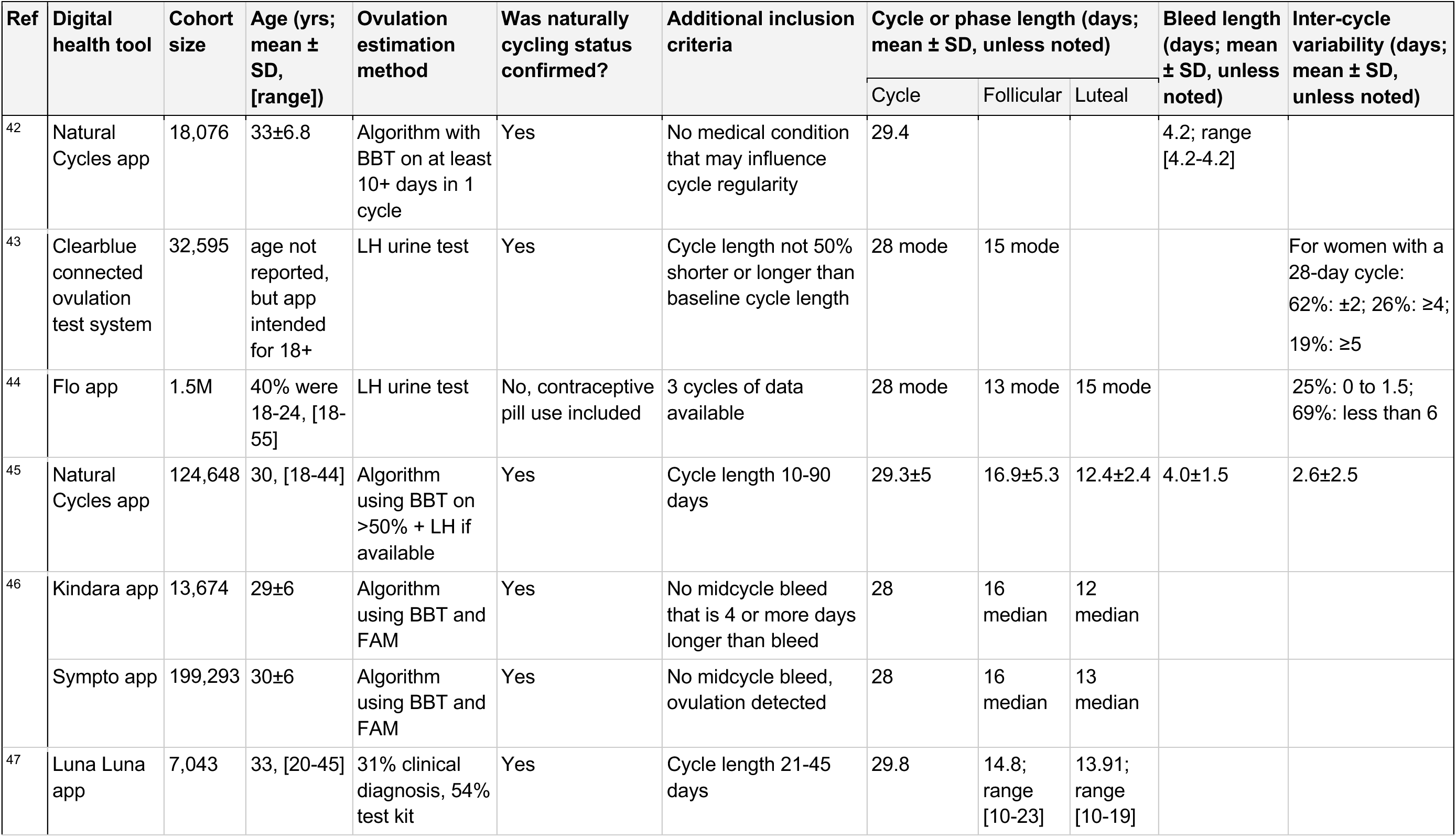
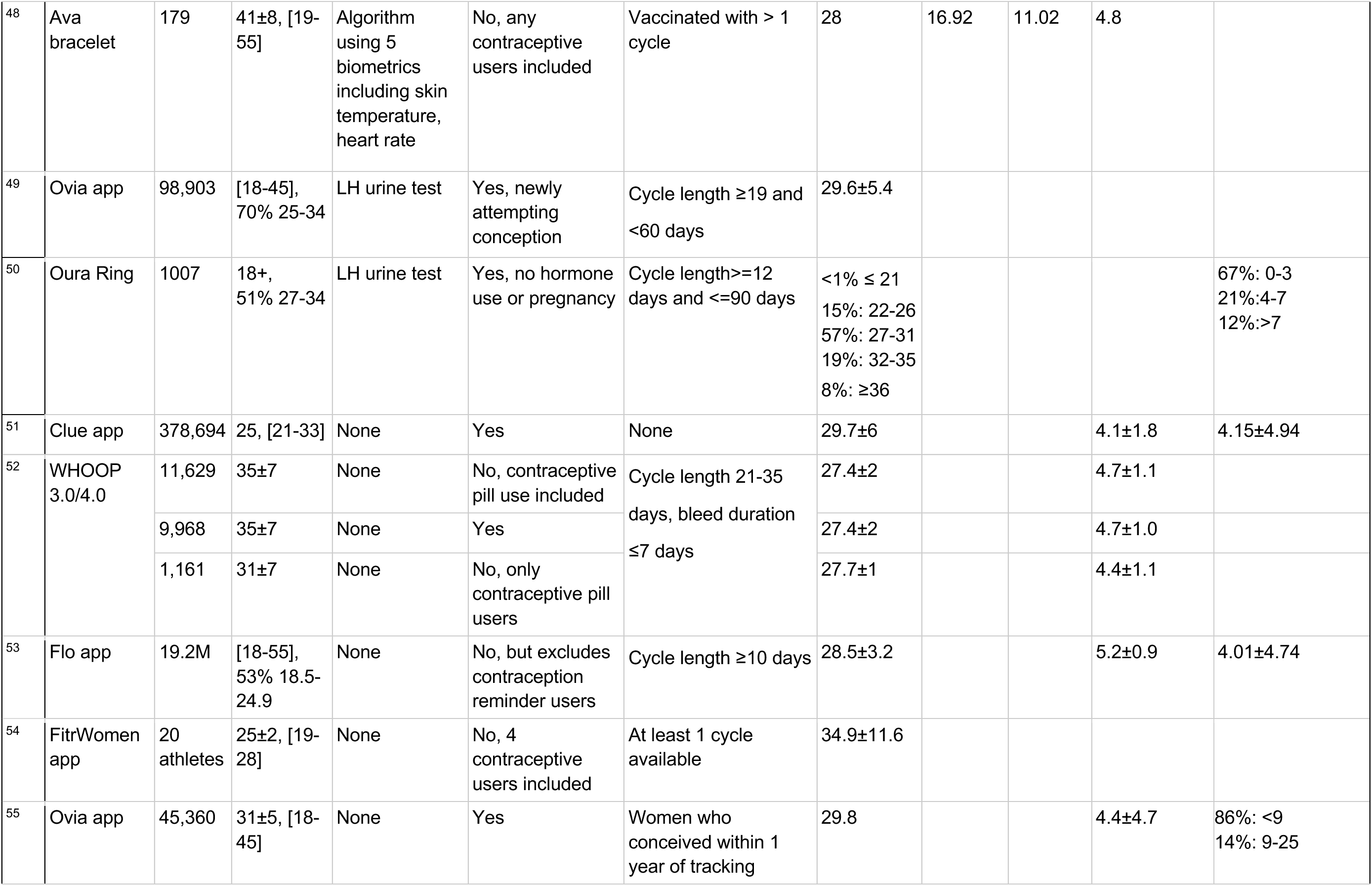
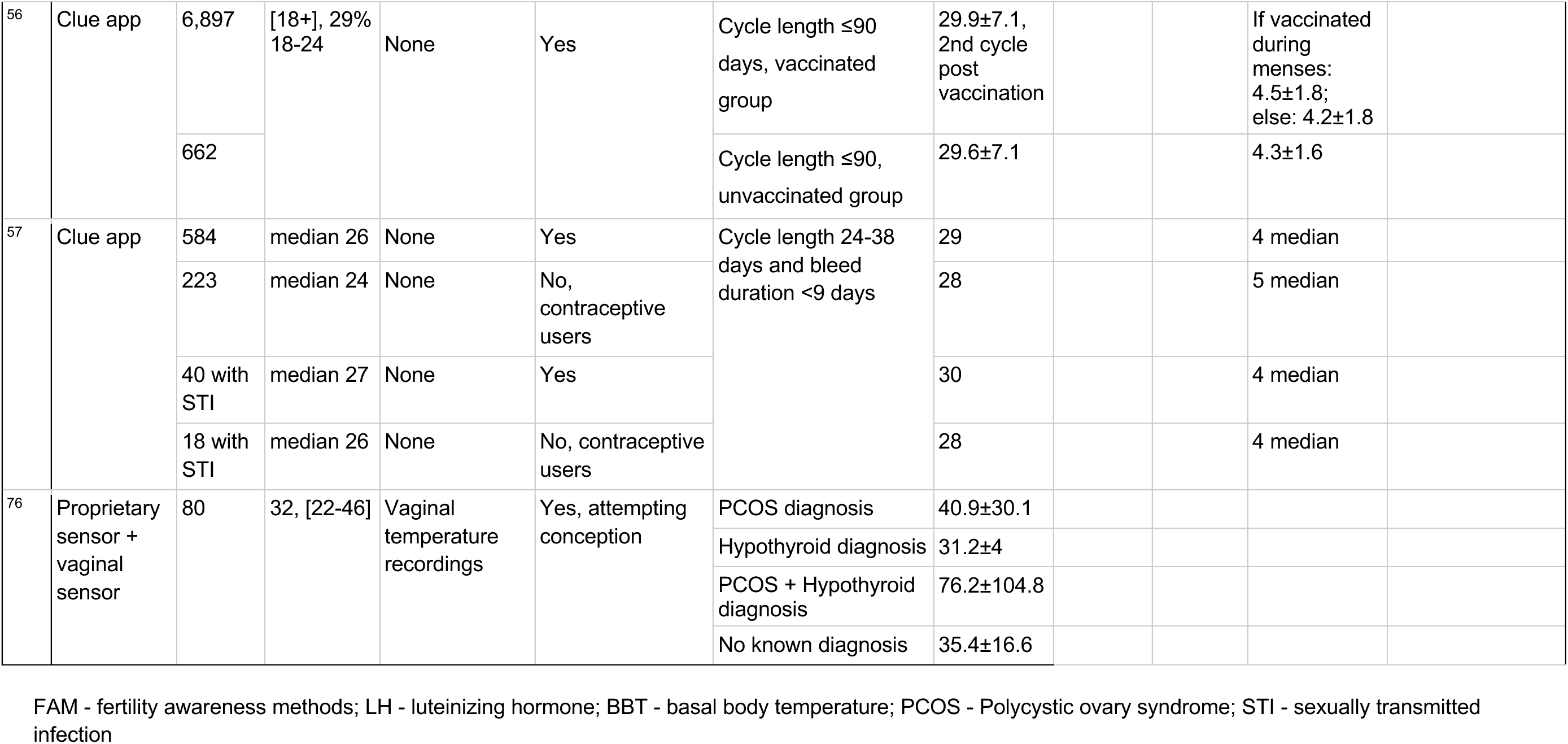
Menstrual cycle characteristics recorded with digital health tools. In this table, we summarise the results of all studies that reported at minimum cycle length and age.

**Table 2:**
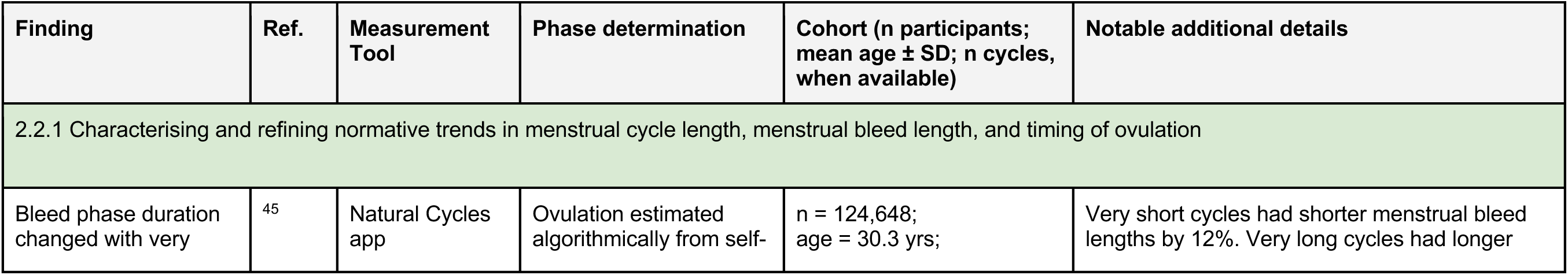
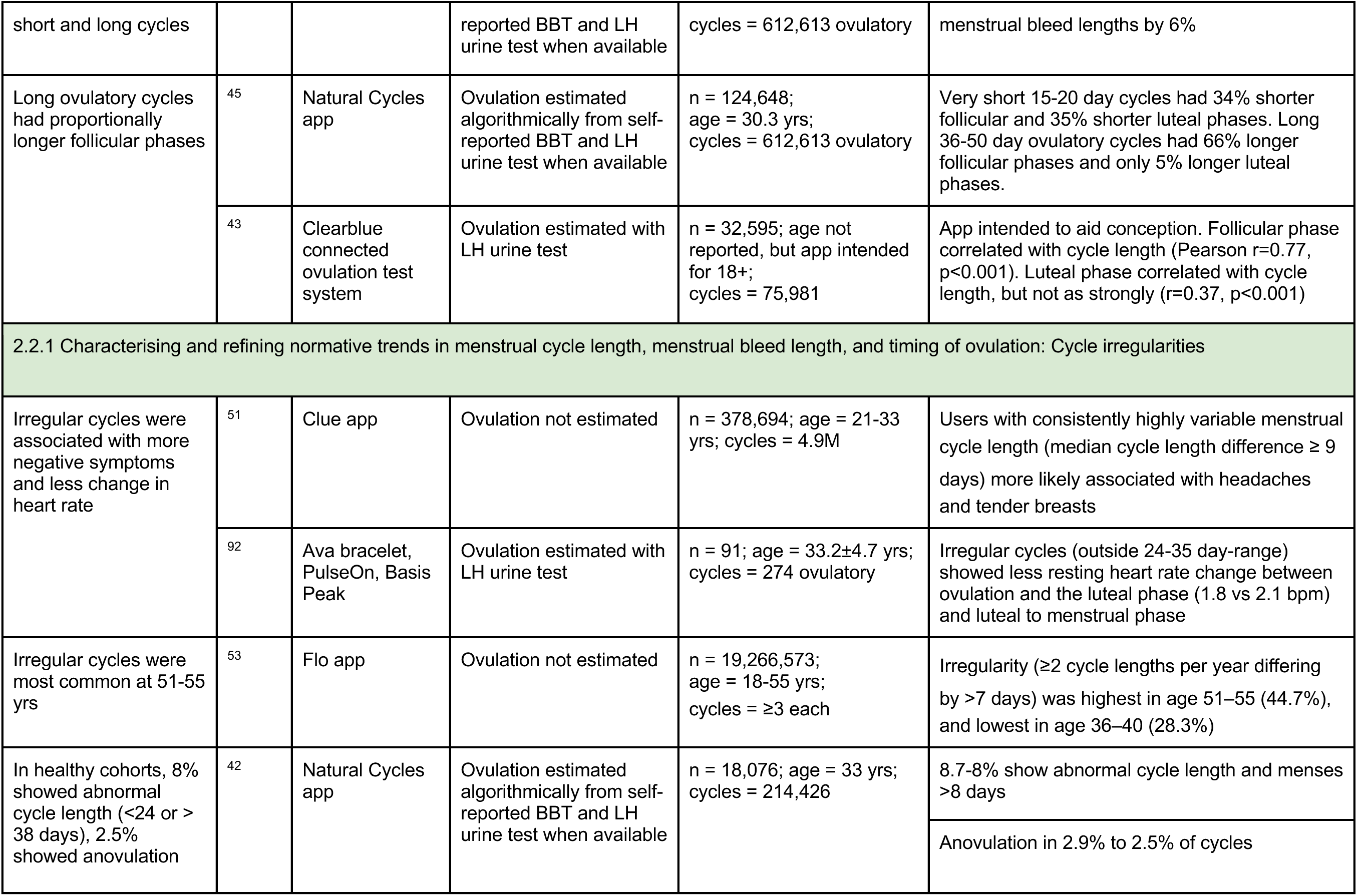
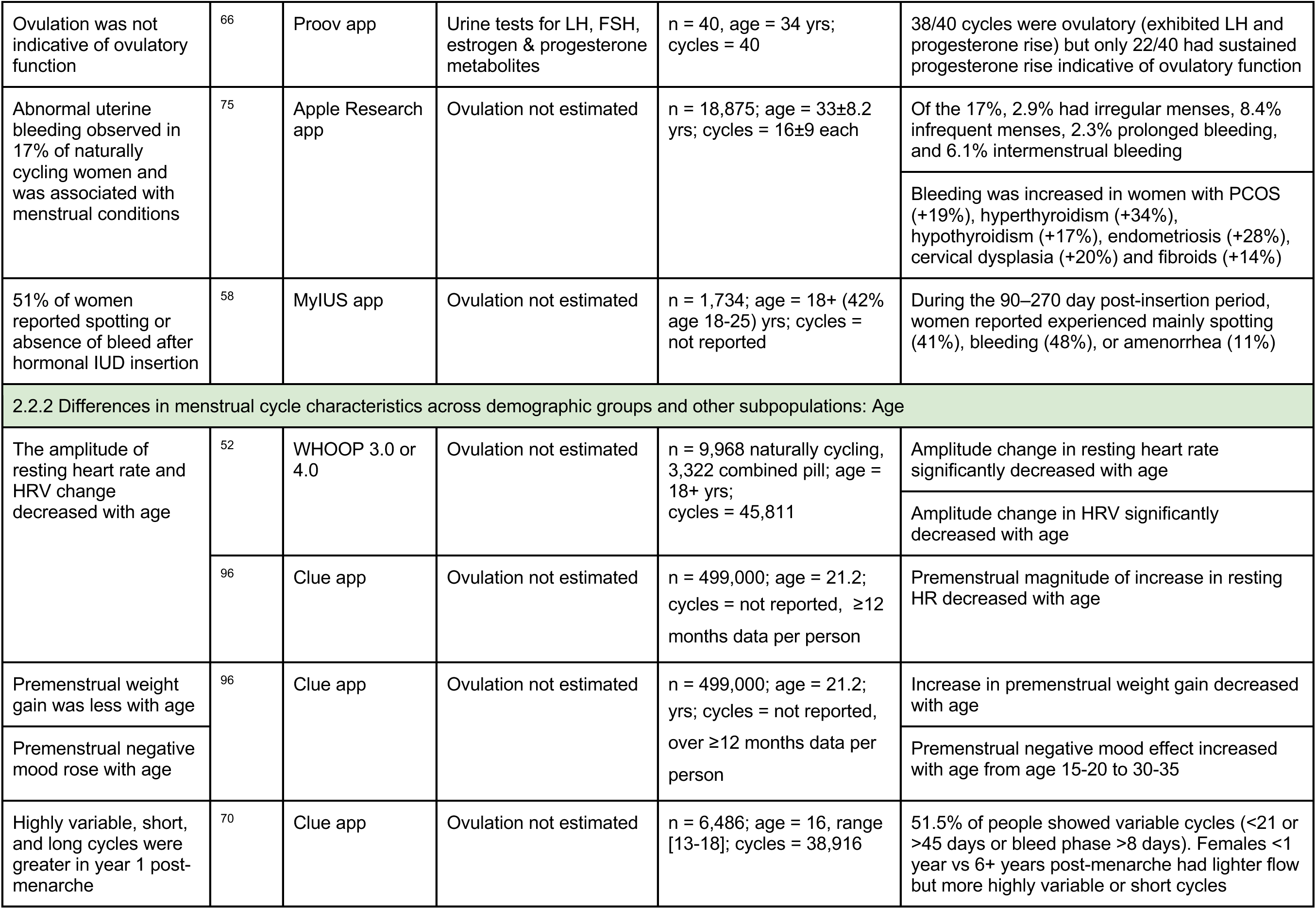
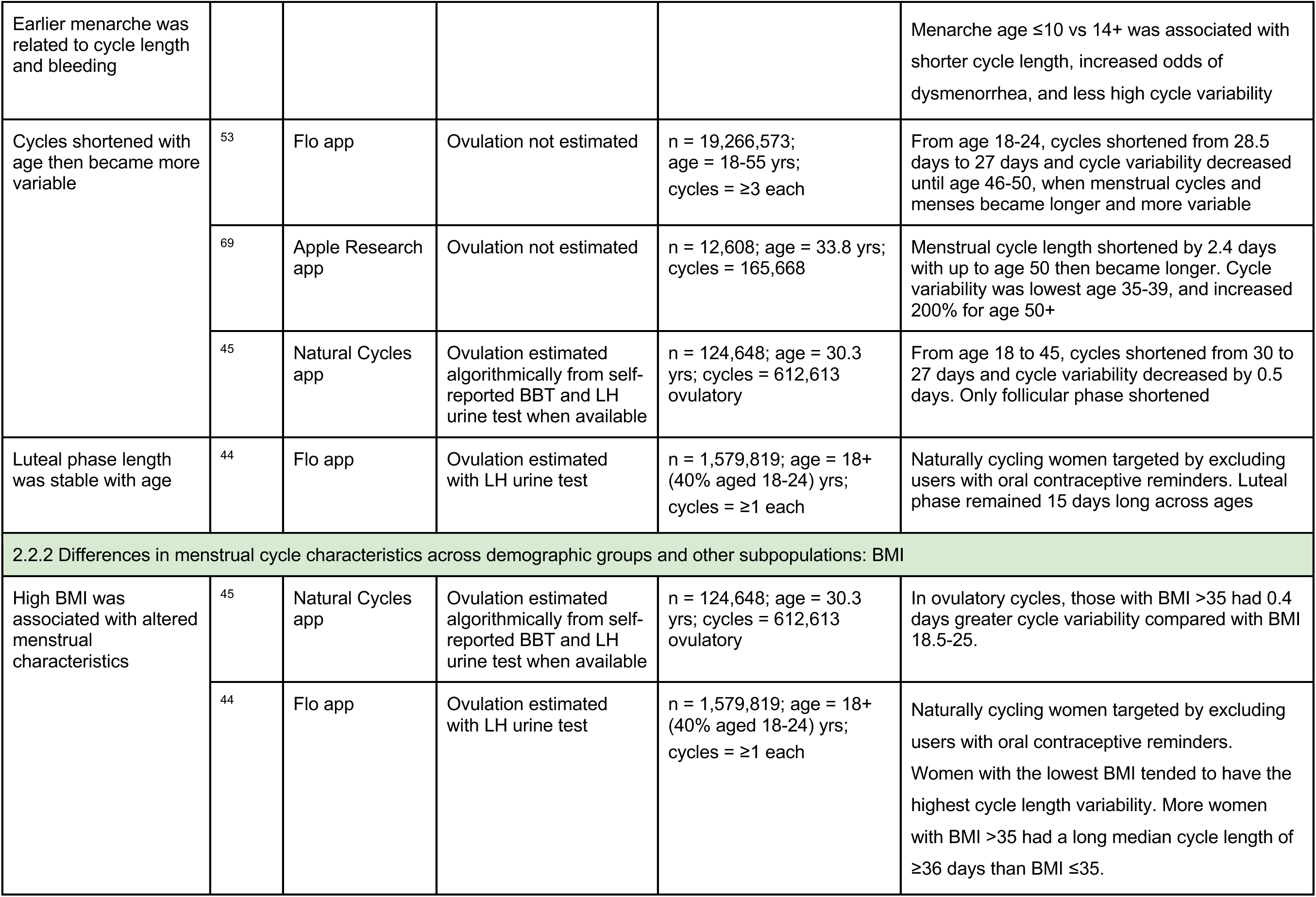
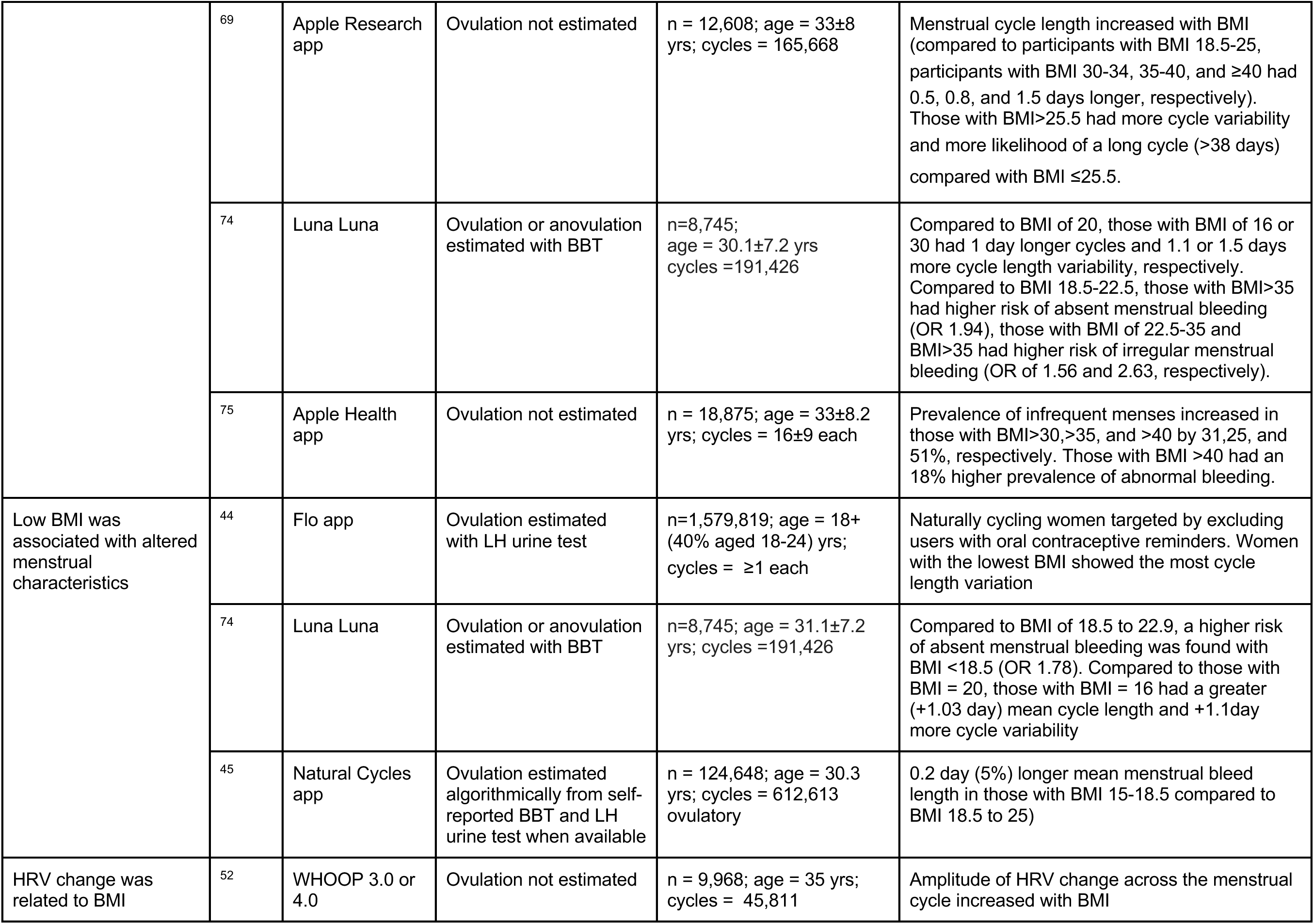
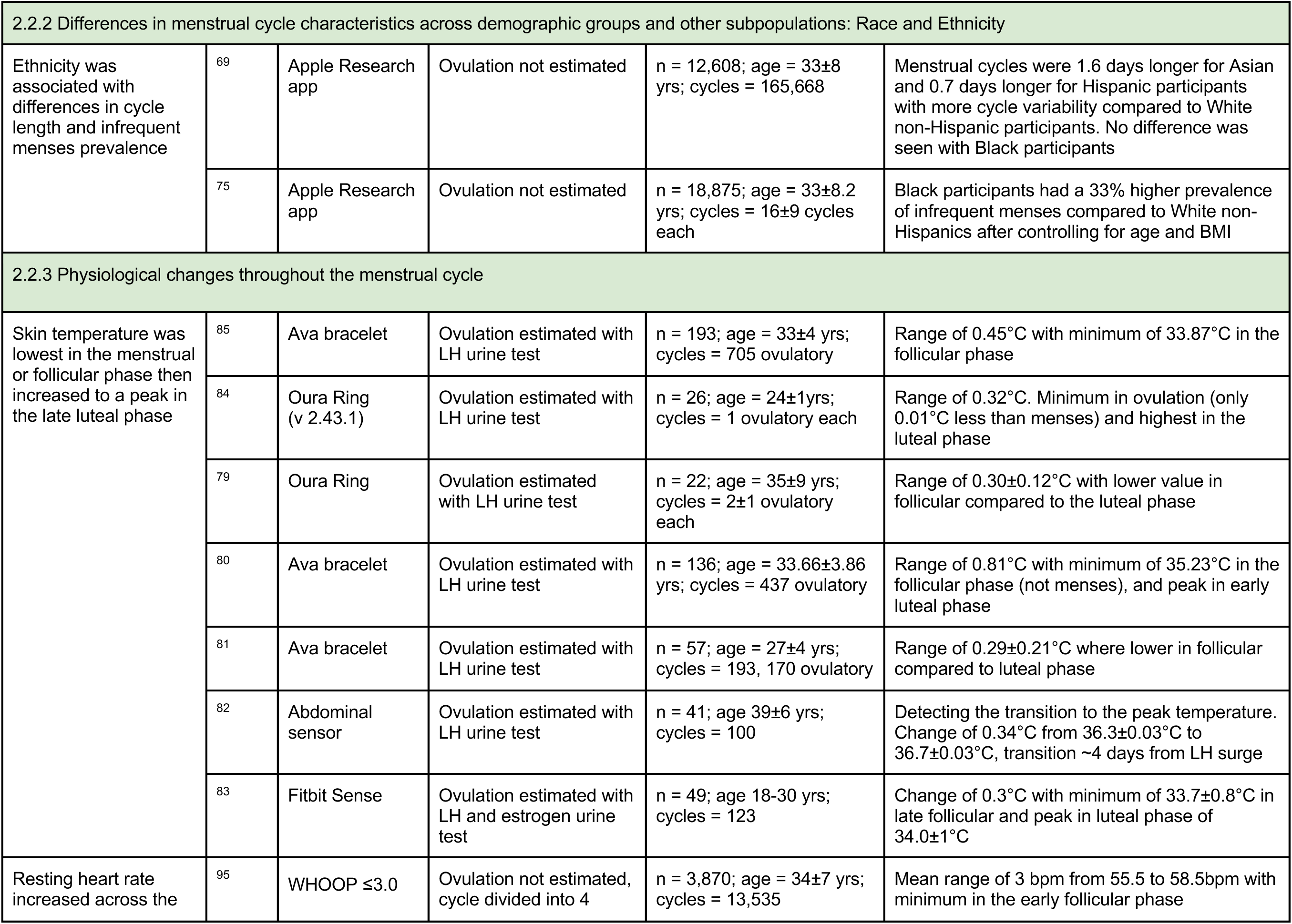
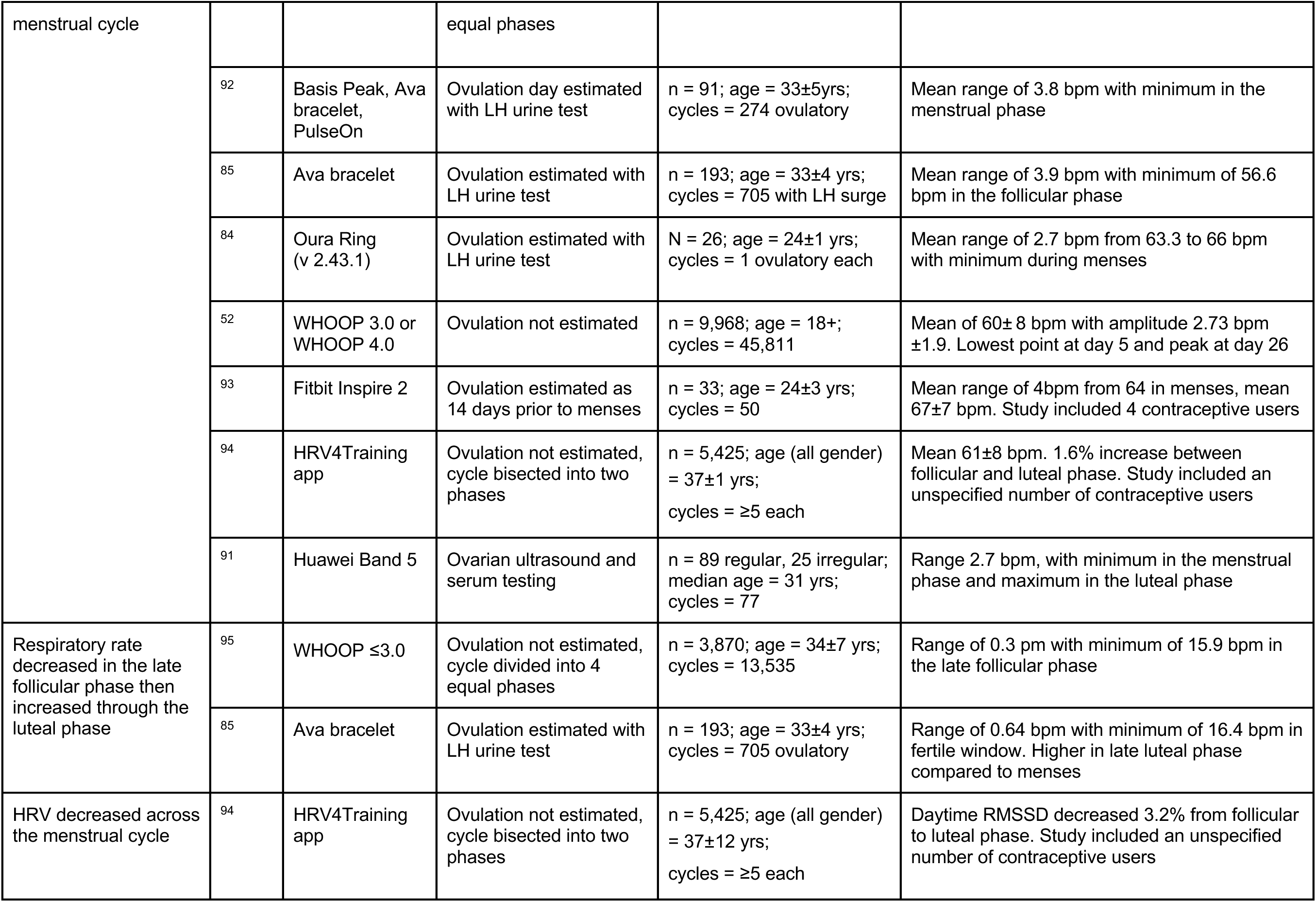
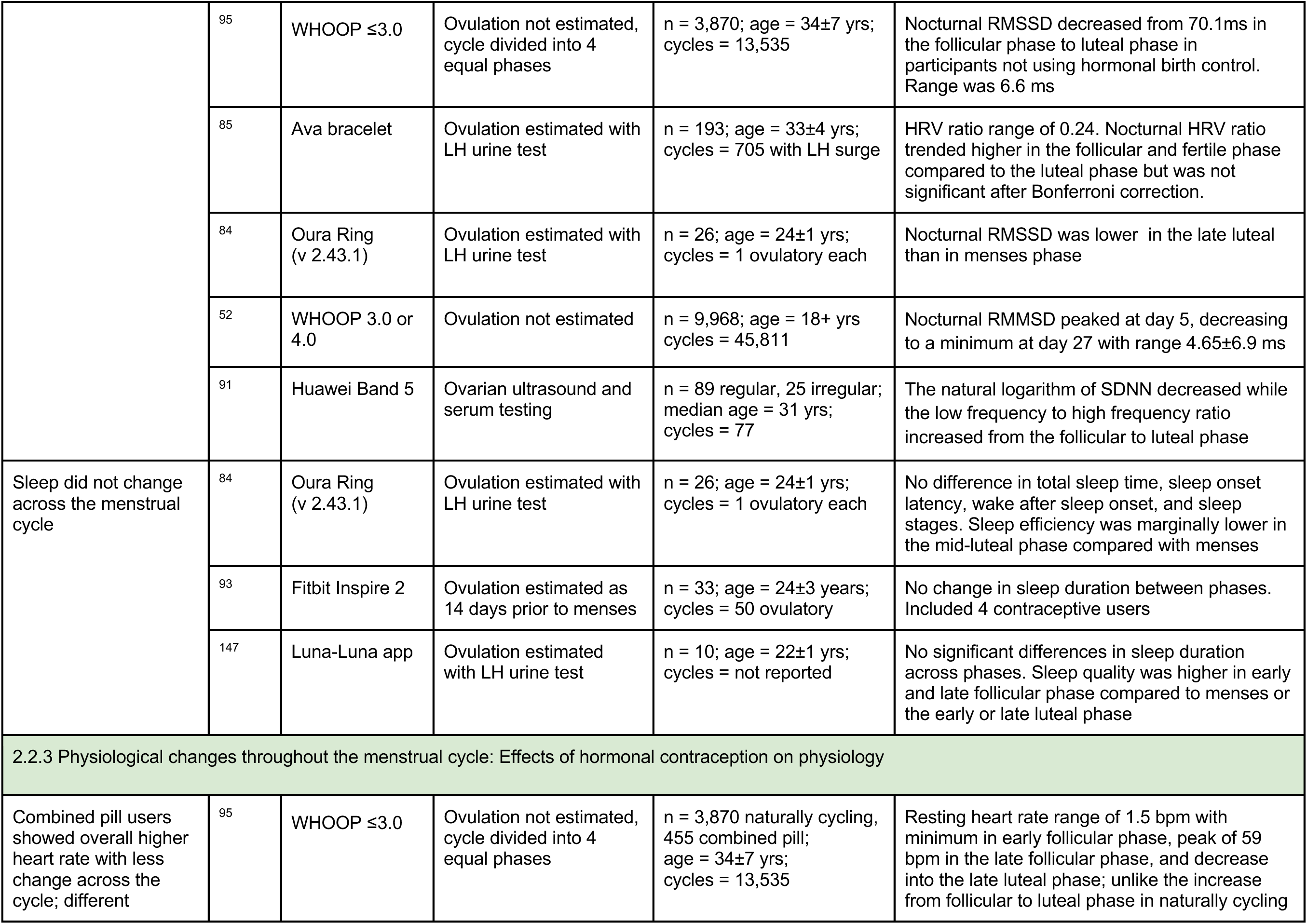
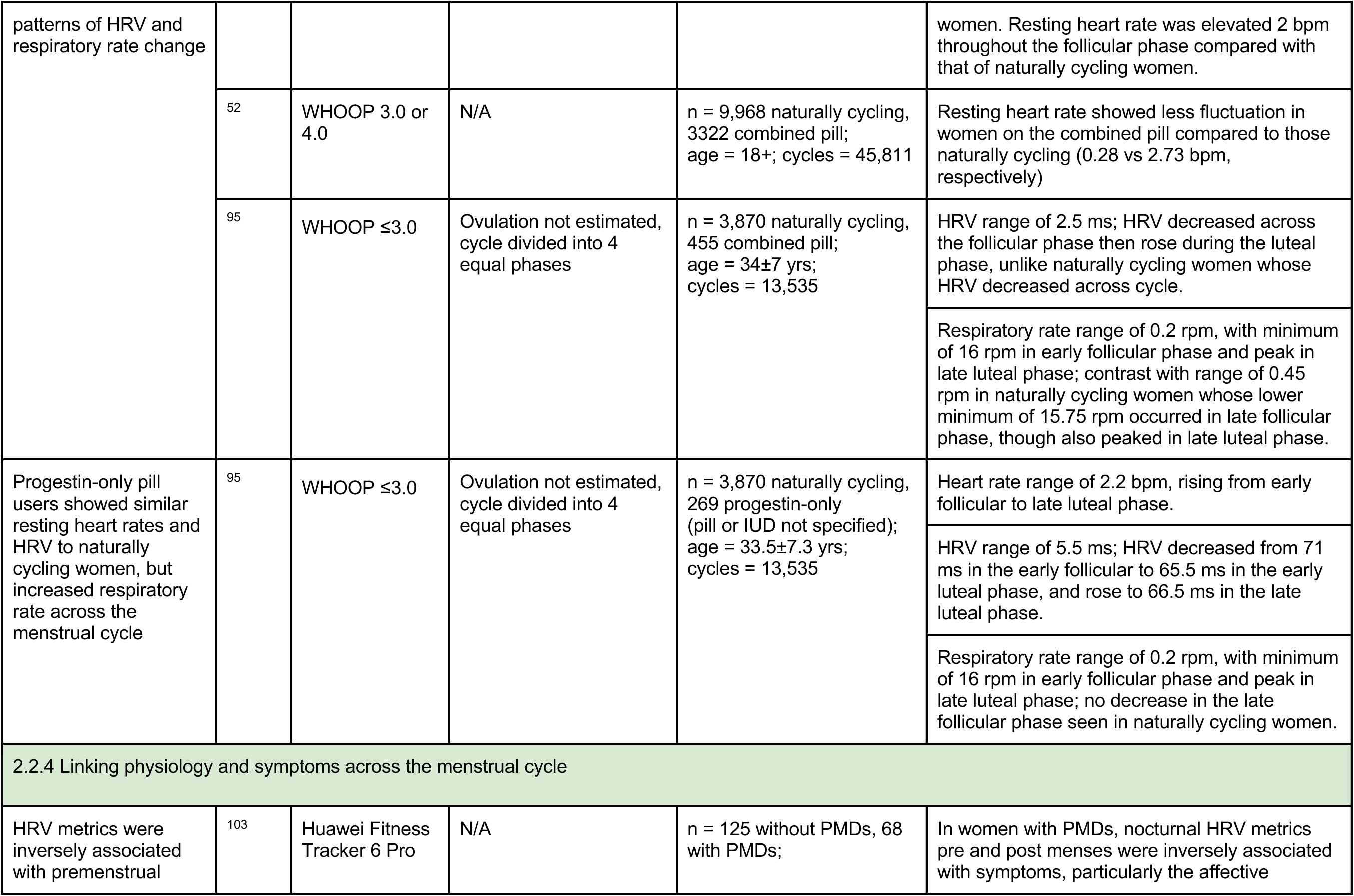
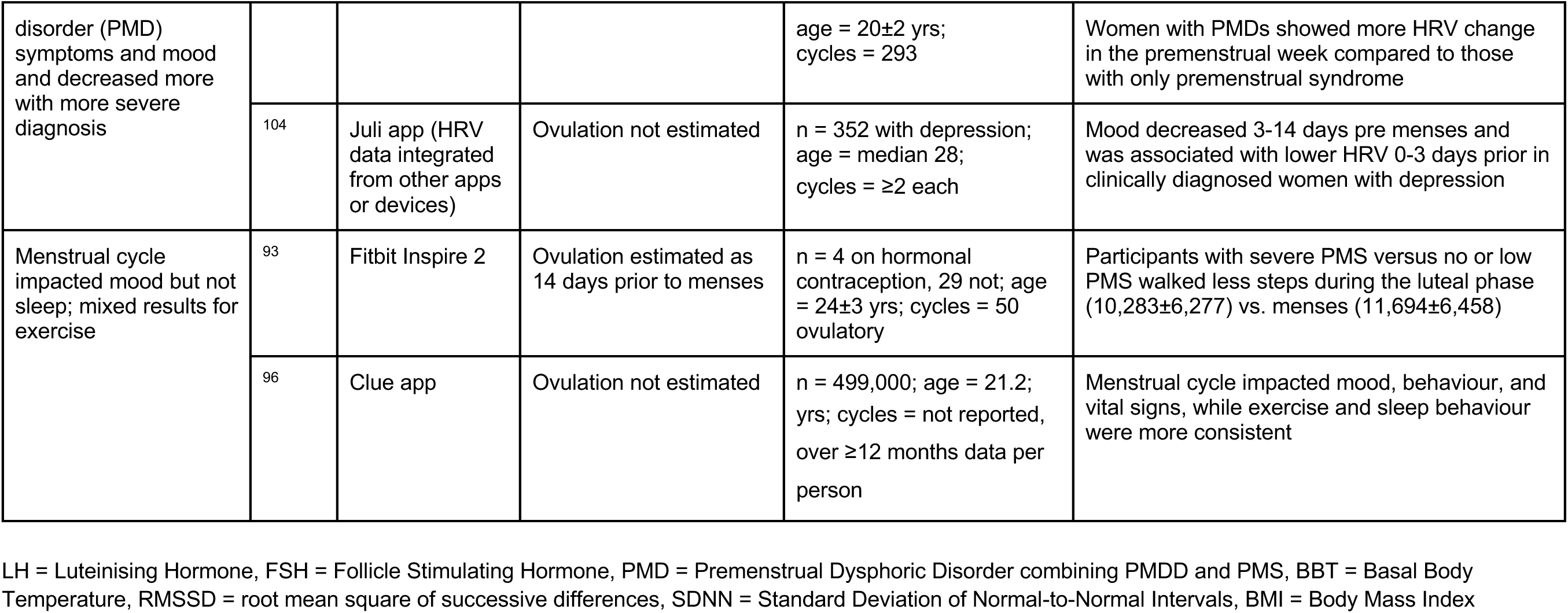
Insights from monitoring of menstrual cycles with digital health tools. Naturally cycling women, unless otherwise stated.

### 2.2 Insight gained into physiology and behaviours accompanying menstruation

Digital health studies have enabled researchers to refine our understanding of how menstrual cycle physiology varies across lifespan. In this section we summarise the reported characteristics of menstrual cycles and the limited emerging research that uses the capabilities of digital health tools to compare menstrual cycle characteristics across demographic groups and other subpopulations, and to connect physiology and menstrual cycle characteristics with symptoms, physical activity, and sleep.

#### 2.2.1 Characterising and refining normative trends in menstrual cycle length, menstrual bleed length, and timing of ovulation

##### Menstrual cycle length and variability

Understanding typical cycle lengths and variability, both within and between women, is crucial to distinguish natural variability from underlying health concerns^39^. Traditionally, a mean menstrual cycle length of 28 days with a 21-35 day range has been considered “normal” and was used as an inclusion criteria in many of the studies^40^. In 2018, the International Federation of Gynecology and Obstetrics (FIGO) guidelines for the range of cycle lengths were updated to 24-38 days^40,41^. Across the majority of studies we reviewed, the average “ovulatory” menstrual cycle determined typically with self-reported LH testing or basal body temperature) was 28-30 days^42–50^, while menstrual cycles analysed without any estimate of whether ovulation occurred averaged 27-30 days^51–57^ (Table 1). The standard deviation of menstrual cycle length between people varied from 2.3-7.2 days and variability within women from cycle to cycle averaged 2.6-4.4 days. However, these values for average length and variability may not be fully representative, since studies often excluded conditions that could impact the menstrual cycle such as hypothyroidism or endometriosis and women with shorter or longer cycles than “normal”. The general agreement in average cycle length between digital health studies and prior work provides confidence in using digital health technology to track cycle length in large and free-living populations, but additional studies with broader inclusion criteria are needed to better characterise the full spectrum of menstrual cycle characteristics.

Indeed, three studies, each including over 10,000 women reported that between 8% and 13% of women had ovulatory menstrual cycle durations outside “normal” ranges (i.e., fewer than 21-24 days or more than 35-38 days), depending on the study, again with ovulation estimated from LH urine testing or basal body temperature^42–44^. Furthermore, in a study of 378,000 women, 8% of the women showed median cycle-to-cycle variability of 9 days^51^, and three studies reported that over 19% of women had a greater than 4^50^ and 5 day cycle-to-cycle variability^43,44^.

Meanwhile, women using any type of contraceptive pill that suppresses endogenous hormones showed no cycle length changes compared to those naturally cycling, (cycle length = 27.7±1.44 days)^52^ though this is consistent with a standard 28-day pill regimen. We found one digital health study analysing bleed intensity in the 3 to 9 months following hormonal IUD insertion and found that 10% had amenorrhea and 41% had irregular cycles due to spotting (fewer than 8 total days of menstrual bleeding over the 6 months)^58^, underscoring the disruption of menstrual cycle characteristics. Digital health tools are uniquely positioned to validate preliminary evidence linking progestin dosage to amenorrhea rates^59,60^ and to explore dose versus endogenous hormones patterns via remote urine test logging. Large-scale digital health studies have also enabled researchers to begin to analyse variations in cycle length and other characteristics across demographic groups and other subpopulations (see Section 2.2.2), and connect cycle characteristics with symptoms (see Section 2.2.4).

##### Menstrual bleed length

Abnormal uterine bleeding can indicate underlying health problems and 4.5-8 days is considered a clinically normal menstrual bleed length^40,41^. The studies we reviewed reported a mean or median 4-5 day menstrual bleed length (Table 1), regardless of whether ovulation was estimated or if cycles with prolonged bleeding (over 7 days) were included. A study of over 1,000 participants comparing women on any type of contraceptive pill to those naturally cycling showed no change in menstrual bleed length (4.36±1.09 days)^52^. Menstrual bleed lengths from digital health studies were within the clinically normal bleed length, giving confidence in self-reporting of menstrual bleed length.

##### Phase length and ovulation

Accurately identifying ovulation is crucial for planning or avoiding pregnancy. If sufficiently accurate, digital health tools could improve our understanding of normal inter- and intra-individual physiological variation and be applied to guide conception or contraception.

Identifying changes in phase length can also help to identify shortened luteal phases that may represent insufficient progesterone production that can impact fertility and/or be an indicator of low energy availability in athletes^61–63^. Data collected with digital health tools show that ovulation may occur later in the cycle than the previously expected day 14^64^ and with more variable timing, with one study reporting a 10-day range in ovulation day for women with a 28 day cycle^43^. Five out of the six studies that estimated ovulation and reported the mean, median or mode follicular phase length (Table 1)^43,45–48^ reported a longer length of 15-17 days, compared to 13-14 days from lab-based studies^65^.

The method of ovulation method detection varied between studies, with two studies reporting ovulation via self-reported LH urine test results^43,44^, one study also accepting any type of clinical diagnosis^47^, two studies using algorithms based on self-reported basal body temperature and symptoms^46,48^, and one study using an algorithm based on self-reported basal body temperature and LH urine test results, where available^45^. The variation between studies in cycle length inclusion criteria, measure of central tendency, and ovulation estimation approach could all contribute to the differences between studies. For instance, the mean follicular phase length ranged from 15 to 17 days while mode ranged from 13 to 15, suggesting a distribution that is skewed by longer cycles. And studies that incorporated LH urine tests or clinical diagnosis tended to estimate slightly shorter follicular phases than those that relied on basal body temperature and other symptom reporting. Greater rigor and consistency of reporting in future studies, along with a careful meta-analysis of prior studies, could help to resolve our understanding of the mean and range of follicular phase length across a range of demographic and other groups. One study utilised an app-integrated device to monitor four hormones, enabling potential future studies to confirm ovulation through the dual detection of LH surges and progesterone^66^.

Lab-based studies have reported greater variability in the length of the follicular phase compared to the luteal phase, which suggests that an extended follicular phase may be the primary driver of longer menstrual cycle length^65,67^. Studies using digital health tools support this finding, with a strong correlation between overall menstrual cycle length and the follicular phase (r=0.77), but only a weak correlation with luteal phase (r=0.37)^43,45^. Furthermore, in short menstrual cycles, the follicular to luteal phase ratio stayed constant, but in long ovulatory menstrual cycles (36-50 days) the follicular phase lengthened by 66% with a minimal luteal phase increase (5%)^43,45^. The luteal phase length stayed consistent across age groups despite menstrual cycle length changing with age (see Section 2.2.2), providing additional support for the concept that ovulation timing drives cycle length^43,44^.

#### 2.2.2 Differences in menstrual cycle characteristics across demographic groups and other subpopulations

##### Age

Studies using self-reported cycle tracking apps (with or without ovulation tests) support prior findings that menstrual cycle characteristics shift with age. In line with lab-based studies^68^, for individuals from age 18 to 25, digital health studies showed highly variable menstrual cycle lengths^45,53^. After age 25, cycle length shortened with increasing age due to an estimated shortening of the follicular phase^44,45,53^. When comparing across the study populations, the standard deviation for menstrual cycle length remained similar from age 18 to 30, then increased for individuals over age 30^45^ and 40^53,69^.

Digital health studies have also found new relationships between age of menarche and cycle length and symptoms. A study using data reported by adolescents in the United States aged 13-18 who used the Clue app found that mean menstrual cycle length was longer (32.4±6.8 days) and the odds of highly variable, short, or long cycles was greater in the first year of menarche compared to six years post menarche^70^. Furthermore, shorter cycles and increased reports of dysmenorrhea were associated with earlier menarche onset^70^.

##### Body Mass Index

BMI is known to impact menstrual cycle characteristics, with low BMI associated with amenorrhea due to hypothalamic suppression^71^. Changes with high BMI have been mixed in prior studies. One study found longer cycle lengths in individuals with high BMIs based on a retrospective review of medical records^72^, while another found no change in cycle lengths, but lower progesterone, LH, and FSH and higher free estrogen in individuals with BMI of 30 and above^73^.

Digital health studies have shown that cycle length, cycle length variability, abnormal bleeding, and absent or infrequent menstrual cycles are increased for both low (under 18.5) and high (over 25) BMI, though underweight cohorts remain underrepresented (2-9%). Compared to those with a BMI between 18.5 and 25, those with low BMI showed the highest cycle length variability^74^, reported slightly longer menstrual bleed lengths (0.2 days increase)^45^, and longer cycles^74^. Meanwhile at high BMI, longer cycles^69,74^, increased cycle variability^45,69,74^, and increased abnormal uterine bleeding^74,75^ were reported in some but not all studies. Some of these effects were small, for example cycle length increased on average by 0.5 and 1.5 days with BMI above 25 and 40 respectively^51^. Discrepancies between studies, such as the lack of change in cycle length in those with high BMI reported by Bull et al.^45^, may stem from the exclusion of anovulatory cycles. In contrast, studies like Itoi et al.^74^, that included non-ovulatory cycles may capture a wider range of hormonal disruptions, revealing more significant differences. Further, Itoi was able to find an increased incidence of anovulatory cycles in those with low and high BMI, a finding that warrants further exploration. Together, the larger size of these digital health studies (e.g., including thousands of individuals with high BMI), along with the regular collection of cycle length data (in contrast with cycle details reported to a clinician and stored via medical records) has revealed the valuable new evidence connecting BMI with cycle characteristics and uncovered trends (such as the connection between cycle variability, anovulatory cycles, and both low and high BMI) to explore in further research.

##### Race and ethnicity

We found that most studies reviewed did not report ethnicity, and those that did predominantly included participants from the United States, United Kingdom, and Europe with predominantly White cohorts. Despite the sparse data, the use of digital health tools has begun to show that there may be differences in menstrual characteristics across different racial and ethnic groups. For example, Asian and Hispanic participants reported longer and more variable menstrual cycles^69^, and Black participants showed a 33% higher prevalence of infrequent menses compared to White non-Hispanic women^75^. There is a clear need for research in more diverse populations in order to understand when there may be meaningful differences between groups.

##### Other subpopulations

Digital health studies have also identified that cycle lengths can be longer and more variable in other subpopulations. One study of athletes, which did not exclude participants based on cycle length, reported a longer mean menstrual cycle length of 35 days with a 12-day standard deviation, reflecting the increased incidence of menstrual irregularity in athletic cohorts^54^. There is ample opportunity to examine menstrual cycle characteristics in other groups who may be exposed to physical or other stressors, including shift workers or those with varying socioeconomic backgrounds.

A study of women trying to conceive found that mean cycle length and standard deviation varied dramatically in those with menstrual conditions. Women with polycystic ovary syndrome (PCOS), hypothyroidism, or both conditions reported a mean cycle length of 41, 31, and 76 days respectively^76^. This characterisation of cycle-length differences between cohorts could be a first step toward earlier diagnosis or better monitoring.

#### 2.2.3 Physiological changes throughout the menstrual cycle

Understanding changes in biometrics such as temperature and heart rate is relevant for a variety of health and performance applications. For example, characterising subtle temperature dynamics around ovulation is important because many proprietary algorithms that aim to detect the fertile window in near real time rely on detecting the well-characterised temperature rise around ovulation. Resting heart rate, HRV, and respiratory rate can be indicators of fitness, recovery, and overall wellbeing, while unexplained changes in these biometrics can indicate illness and stress. Establishing baselines for biometric fluctuations across the cycle and lifespan can help women understand whether deviations coincide with changes in health or performance or menstrual cycle irregularities. We found that digital health studies using passive monitoring with wearable sensors have reproduced known trends for skin temperature, resting heart rate, and respiratory rate (Figure 1), providing confidence in the technology’s utility for studying deviations from the norm that may accompany behavioural or health changes. The results have also provided new evidence for HRV fluctuations across the menstrual cycle, improved resolution (i.e., daily rather than weekly) of physiological patterning across the menstrual cycle, and quantified how these trends change with age.

**Figure 1:**
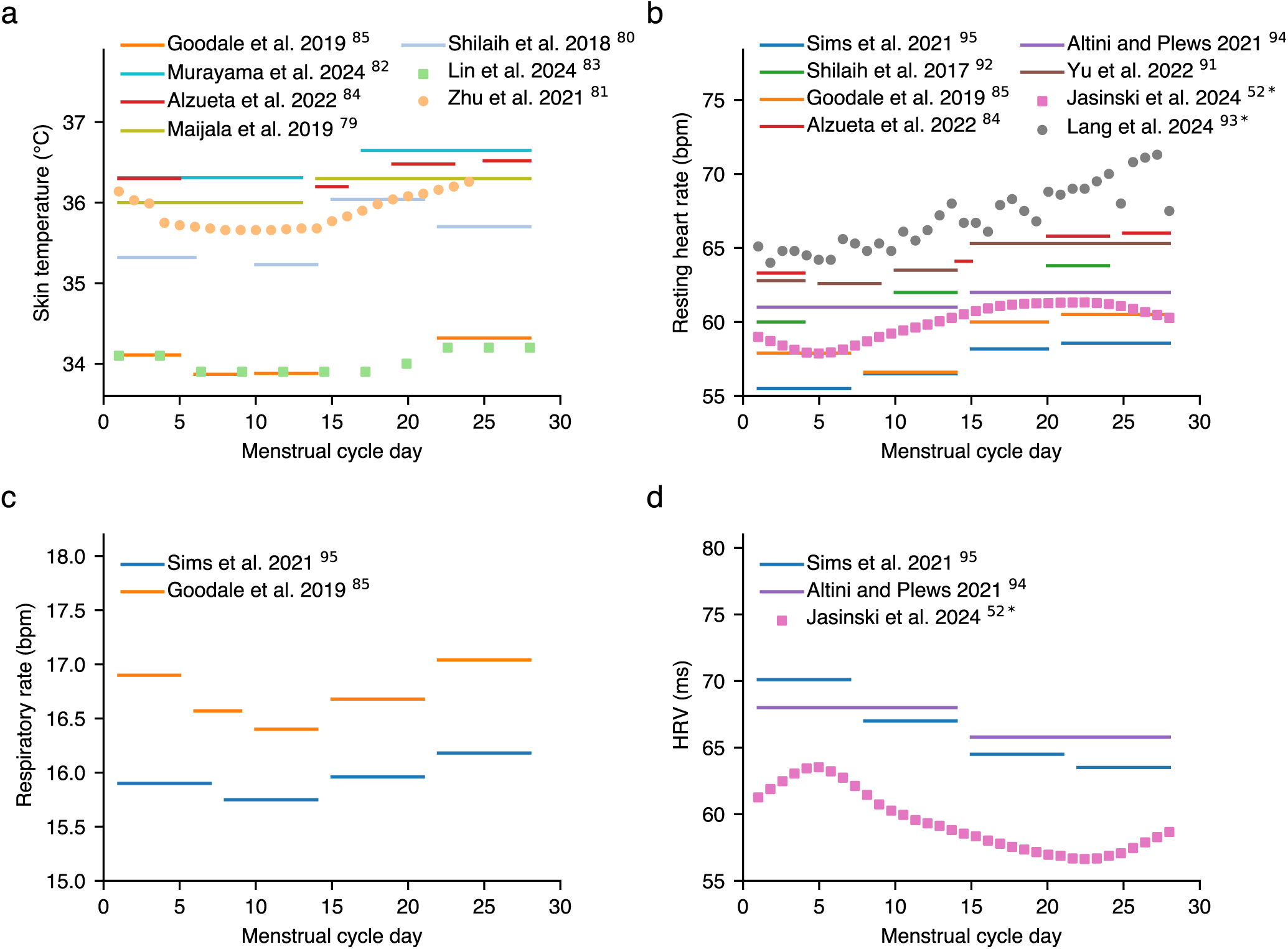
Reported changes in biometrics across the menstrual cycle, as recorded with wearables. Biometrics that have been characterised include a) skin temperature, b) resting heart rate, c) respiratory rate, and d) heart rate variability (HRV). Where phases were estimated based on time from menses or ovulation, we assumed a 28-day cycle with ovulation on day 14. Data from longer cycles (35 days; studies noted with *) were normalised to a 28-day cycle. Resting heart rate is in units of beats per minute. HRV is shown as the root mean square of successive differences (RMSSD) in milliseconds, either directly reported or calculated from reported percentage change in fluctuation from a mean. Respiratory rate is breaths per minute at rest. Skin temperature is in degrees Celsius measured at rest.

**Figure 2:**
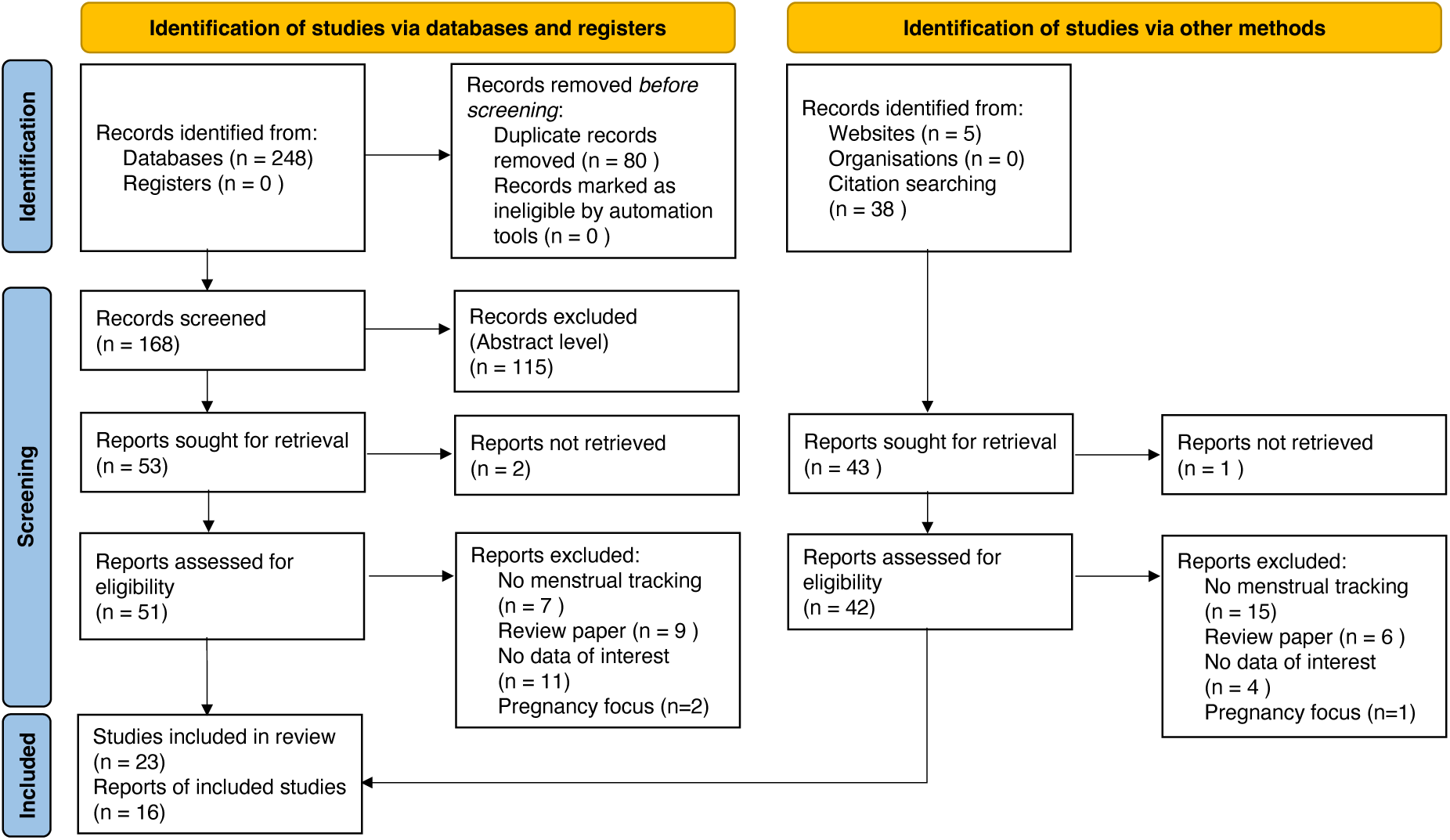
PRISMA flow diagram in the search of papers associated with the menstrual cycle. Adapted from the PRISMA flow diagram^146^.

##### Skin temperature

Laboratory studies show that core body temperature is 0.3-0.7°C lower in the follicular phase than the luteal phase^21,77^ and that the minimum basal body temperature occurs one or two days before ovulation, or around day 12-14 for a typical 28-day menstrual cycle^78^. This pattern of change in temperature is used to predict the fertile window around ovulation which indicates the start of the luteal phase. We found that studies using wearables (ring, wrist and abdominal sensor) reported that skin temperature measurements were similarly 0.3-0.5°C lower in the follicular phase as compared with the luteal phase (Figure 1a)^79–85^.

The continuous nature of wearable-derived biometrics can capture temperature shifts that may be missed by the discrete sampling of lab or clinical studies. Data from wearables showed that skin temperature decreased the most in the first 5 days of the menstrual cycle, in some cases reaching a minimum as early as day 5 then either stabilising until estimated ovulation^81,85^ or declining slightly until a few days pre-ovulation^83^ (Figure 1a). Shilaih et al. found that in 43% of cases the lowest temperature occurred within the five days prior to ovulation, but in 47% of cases the lowest temperature occurred earlier^80^. One exception is that Alzueta et al. reported a minimum temperature at estimated ovulation, but this may be due to a lack of measurements between day 5 of the cycle and ovulation^84^. Taken together, these studies (Table 2) suggest that while most temperature decrease occurs by the end of menses, the specific minimum point before ovulation may vary, although whether due to inter-individual differences, inter-cycle variability, or study methods is unclear. All studies that separated subsequent early and late luteal phases found that skin temperature peaked in the late luteal phase^81,84,85^.

One limitation that could contribute to different results is definitional heterogeneity regarding phases such as ‘pre-menstrual’, ‘fertile days’, or ‘ovulatory days’, which hinders cross-study comparison and underscores the need for standardised terminology. Furthermore, in most studies using wearables and in many lab studies, ovulation was confirmed with LH urine testing alone. While urinary LH testing has been shown to have high accuracy in detecting LH surges that trigger ovulation^86^, with the development of at-home kits to detect progesterone metabolites from the subsequent progesterone release upon egg release^66^, confirmation of ovulation could be further improved and the relationship to skin temperature dynamics further validated and refined.

##### Resting heart rate

Some lab studies reported an increase in heart rate in the mid luteal phase^87,88^; one found no change across the menstrual cycle^89^, and another reported an increase at ovulation only^90^, though all had cohorts on the order of 10s of participants recording heart rates at discrete time points. Digital health studies use frequent, passive measurement of heart rate, usually during sleep. While cohort sizes have ranged from smaller pilot groups^84,91^ to large-scale population studies exceeding 10,000 participants^52^ they have consistently shown that average resting heart rate increased from the follicular phase to the luteal phase in naturally cycling women by 2.7-3.9 bpm (Figure 1b)^52,84,85,91–95^. Four of the studies with more granular data found that the largest change in resting heart rate occurred mid-cycle, specifically in the 5 days leading up to, and including ovulation when compared to post ovulation. Two studies with nearly 510k participants additionally report that the amplitude of the change in resting heart rate across the cycle becomes smaller with age^52,96^.

##### Respiratory rate

Digital health studies have shown that respiratory rate changed between 0.3-0.6 breaths per minute across the menstrual cycle, decreasing from menses to the pre-ovulation days, then increasing and peaking in the late luteal phase (Figure 1c)^85,95^. This higher luteal phase respiratory rate is in accordance with past laboratory-based studies showing resting respiratory rate increased by 0.62 breaths per minute in the luteal phase^97^.

##### Heart rate variability (HRV)

A systematic meta-analysis of 37 lab-based studies reported that, despite some contradictory findings, overall, HRV-associated measures in the follicular phase were significantly higher than those in the luteal phase^98^. Similarly, digital health studies have consistently found a higher HRV in the first half of the menstrual cycle (Figure 1d). These studies totaling over 19,000 participants also found that HRV continued to decrease across the second half of the menstrual cycle, with the lowest HRV in the late luteal phase^52,84,85,94,95^. Digital health studies differed slightly in their conclusions around the timing of peak HRV. In two studies totaling nearly 14k participants, Sims et al. found HRV peaked in the early follicular phase (including menses)^95^, while Jasinski et al. reported peak HRV the day after menses (day 5)^52^. In contrast, a smaller study of nearly 200 women by Goodale reported phase-based differences that only trended towards significance^85^. Jasinski et al.^52^ also found that when using a generalised linear model, the amplitude of HRV change between phases decreased significantly with age, mirroring their findings on age-related decline in resting heart rate amplitude. The variation in peak timing could be due to the accuracy of sensors (see Section 2.4), definition and resolution of menstrual phases, and also mixed evidence that suggests HRV is affected by psychological health^99^.

#### Effects of hormonal contraception on physiology

Use of different types of hormonal contraception can alter the physiological patterns observed throughout a menstrual cycle, which may influence contraceptive choice, particularly in athletes who may track their heart rate or women optimising recovery and monitoring their HRV. Small lab studies (with 10 participants or fewer) have shown use of the combined pill (containing both estrogen and progestin) suppresses endogenous hormones, which in turn flattens the typical biphasic temperature and resting heart rate curves by keeping them permanently elevated^100,101^.

Digital health tools have allowed researchers to begin to characterise these patterns in large cohorts, in greater detail, and with additional types of hormonal contraception. In one study, wrist-worn wearable data from over 450 women using the combined pill showed they exhibited a 2 bpm higher average resting heart rate throughout the follicular phase compared to the naturally cycling women^95^. This elevated heart rate peaked in the late follicular phase at around 59 bpm and then declined through the rest of the cycle (for a total change in magnitude of 1.5 bpm), the inverse of the trend and also over half the range observed in naturally cycling women. HRV fluctuations were also different in those using the combined contraceptive pill. HRV was lowest in the late follicular phase, then rose through the remainder of the menstrual cycle. This is in contrast with the decrease in HRV during the luteal phase in naturally cycling women.

Despite the differences in cardiac biometrics, respiratory rate increased across the menstrual cycle in combined contraceptive pill users, just as observed in naturally cycling women. A second study with a larger cohort of over 3000 women taking any type of contraceptive pill similarly showed less fluctuation in resting heart rate and HRV across the menstrual cycle compared to naturally cycling women (a range of 0.28 vs 2.73 bpm, and 0.51 vs 4.65 ms, respectively)^52^. A separate cohort of the first study focused on over 250 women using progestin-only contraception (either oral or intrauterine), showed overall similar trends in HRV, respiratory rate, and resting heart rate to naturally cycling women^95^. In the luteal phase, HRV was slightly elevated and both HRV and respiratory rate returned to baseline levels more slowly. We did not find any studies that reported biometrics for women using specifically hormonal intrauterine devices or other hormonal forms of birth control.

The lack of delineation in several of these studies between combined pill, progestin-only pill, and hormonal IUD users limits our ability to synthesise and interpret the results and is important to address in future studies. There is a clear demand for women to know what the different effects, if any, that each type of contraceptive has on biometrics and menstrual-related symptoms. The impact of contraceptives on biometrics is particularly interesting as the combined pill can be prescribed to ‘even out’ hormonal fluctuations in women with PMDD yet there remains a lack of studies that have examined the direct impact of contraception on both biometrics and menstrual or premenstrual symptoms—a gap that digital health tools have the potential to fill.

#### 2.2.4 Linking physiology and symptoms across the menstrual cycle

Identifying associations between biometrics and menstrual-related symptoms could increase our understanding of why symptoms occur and aid in both designing and testing the efficacy of interventions. In lab studies using electrocardiograms each with fewer than 70 women, results are mixed. Lower HRV was reported in women with the most severe premenstrual symptoms during the late luteal^25,26^, luteal phase^24^, the follicular phase^27^, or across the cycle compared to controls^25^, while one study reported a lack of reduction in the midluteal phase compared to controls^102^. There is limited research using digital health tools that assess symptoms and physiology together. However, initial studies with more than a hundred participants, have shown, that HRV was inversely associated with self-reported symptoms in women who met the definition of PMS or PMDD^103^ and with a decline in premenstrual mood symptoms^104^, highlighting the benefit of easier access to larger cohorts.

Changes in menstrual cycle characteristics have also been associated with both symptoms and biometrics. A study assessing 5 million self-reported menstrual cycles found that a cohort of women with consistently high menstrual cycle length variability were more likely to report negative symptoms, specifically heavier bleeds and increased likelihood of tracking headaches and breast tenderness^51^. Another study using a wrist-worn wearable showed that while both women with irregular and regular menstrual cycles experienced elevated resting heart rate during the ovulatory phase compared to the menstrual phase, the increase in resting heart rate was somewhat smaller in women with irregular menstrual cycles (1.8 bpm) compared to regular menstrual cycles (2.1 bpm)^92^. These initial links between symptoms, menstrual cycle characteristics, and biometrics suggest that digital tools could be a future avenue to screen for and monitor conditions such as PMS and PMDD, but much more work is needed to validate whether the changes measured by wearables correspond to hormonal profiles measured with gold-standard approaches.

#### 2.2.5 Sleep and physical activity across the menstrual cycle

Understanding how sleep and physical activity can interact with the menstrual cycle is of interest for health and performance, but few studies exist to-date. Although some women self-report sleep disturbances^105,106^, most studies using polysomnography (an in-lab test that records brain waves and movement) showed little to no change in sleep measures^107^. Only one study in our scoping review assessed sleep with a commercial wearable (a smart ring) across the entire menstrual cycle, revealing that sleep efficiency was marginally lower in the mid-luteal phase compared to menses^84^. This effect, from a study of fewer than 20 women, was consistent with another including over 160 women aged 48 to 59, where sleep efficiency declined over the cycle, plunging in the premenstrual week, as estimated from movement-sensing actigraphy^108^.

Taken together, digital health tools have only been able to detect small changes in sleep efficiency across the menstrual cycle, which is perhaps unsurprising with the still limited accuracy of sleep metrics from wearable devices (see Section 2.4.1), and the undetermined relationships between sleep and reproductive hormones^109^. The three sleep studies reported had cohorts consisting of between 19 and 163 women highlighting an opportunity for insight into relationships between sleep, menstrual cycle, and biometrics from large-scale sleep studies with wearable devices. Furthermore, we did not find any studies assessing the impact of physical activity on biometrics or menstrual cycle characteristics, an open and key area to improve understanding of how behaviour impacts menstrual health.

### 2.3 Insight gained into physiology and behaviours accompanying perimenopause

Overall we found scant passively recorded biometric or behavioural data during perimenopause and limited data regarding change in menstrual cycle characteristics. This is in part because many of the digital health studies we reviewed set inclusion criteria for menstrual cycle regularity that removed the irregular cycles that occur around perimenopause. The studies we found showed that cycle length increased and cycle variability dramatically increased with age^51,53^. Li et al found that compared to 35-39 year olds, cycle variability increased by 46% in a 45-49 year-old-age group and by 200% at age 51-55^51^. Furthermore, Cunningham reported that over half of women reported irregular menstrual cycles by age 51-55^53^.

Markovic et al., who included participants aged 19 to 90 without exclusions based on cycle length or age, found that perimenopausal and menstruating participants had a higher skin temperature than the non-menstruating group (which included 577 postmenopausal women, 2 pregnant, and 223 non-menstruating females)^48^. The menstruating participants also had a higher respiratory rate than the non-menstruating participants. Finally, Costeira et al. developed an app to investigate the role of estrogen, which is considered to be immune stimulatory, in COVID protection and found that premenopausal women, especially those using contraceptive pills, exhibited higher COVID protection compared to postmenopausal women^110^. However, the same protective effect of estrogen was not seen in postmenopausal women on hormone replacement therapy^110^. Thus, the menopausal transition is ripe with opportunity to better understand how ovarian hormones relate to changing biometrics.

### 2.4 Guide to wearables, measured metrics, and associated accuracy

Our literature review revealed good agreement between digital health and lab-based studies, supporting the use of wearables and apps to study menstrual cycle physiology, but continued validation against “gold-standard” techniques is needed. To guide researchers in selecting devices with sufficient accuracy to capture changes in biometrics associated with the menstrual cycle, the following sections summarise studies that directly compare wearables to gold-standard measurement approaches, where available (Table 3). We searched PubMed, Web of Science, Google Scholar, and company websites for studies and public data regarding the accuracy and precision of the commercial wearables used in the digital health studies we found in our scoping review. This included the Oura Ring, WHOOP wristband, Ava bracelet, Garmin watch, Apple Watch, Huawei Band 5, and Fitbit wristband. We provide the reported accuracy of skin temperature, resting heart rate, respiratory rate, HRV, and sleep metrics. To understand the accuracy of wearable devices in capturing physical activity throughout the day we refer readers to prior reviews^111–113^.

**Table 3:** Guide to wearables, measured metrics, and associated accuracy.

| Biometric recorded | Device | Method, sensor type, and sampling/timing of measurement, if known | Accuracy | Reference, study type, N of study |
| --- | --- | --- | --- | --- |
| Skin temp | Oura Ring | Negative Temperature Coefficient sensor; measures every minute | Versus iButton sensor: Accuracy = 0.36°C; Precision = 0.13°C; $r^2 > 0.92$ (real world), $r^2 > 0.99$ (lab) | <sup>116</sup> In house, n=16 (company site) |
| | | | Temperature immediately after wake versus oral temperature: $r = 0.563$, $p < 0.01$ | <sup>79</sup> External, devices provided, n=22 |
|  | WHOOP 4.0 | Temperature sensor during sleep | n/a | <sup>148</sup> (company site) |
|  | Ava bracelet | Temperature sensor | n/a | <sup>149</sup> (company site) |
|  | Apple Watch S8 onwards, all Apple Ultra | Two temperature sensors: one skin facing and one outside-facing to remove environmental bias | n/a | <sup>150</sup> (company site) |
|  | Garmin (Select models only) | Temperature sensor | n/a | <sup>151</sup> (company site) |
|  | Fitbit | Fitbit Sense 1/2 uses a dedicated temperature sensor. Fitbit Charge 4/5/6, Google Pixel Watch 2, Fitbit Inspire 2/3, Fitbit Luxe, and Fitbit Versa use a multi-purpose sensor | n/a | <sup>152</sup> (company site) |
| Resting heart rate | Oura Ring and Oura Ring 2.0 | Blood pulse volume changes are detected through infrared PPG sensors in the ring; measured nocturnally every 10 minutes<sup>153</sup> (company website) | Versus ECG (bpm): $r^2 = 0.996$; Bias = -0.63 | <sup>122</sup> Sponsored, n=49 |
| | | | Versus ECG (bpm): $r^2 = 0.998$; Bias = -0.53, $p < 0.001$; Range: 48-66 | <sup>123</sup> In-house, n=10 |
| | | | Versus ECG (bpm): Absolute bias = $1.8 \pm 4.1$; Bias = $0.1 \pm 4.5$; Intraclass correlation = 0.85 | <sup>121</sup> External, n=53 |
| | WHOOP 3.0 | PPG sensor in non-wake in the primary sleep episode, every 30 seconds<sup>154</sup> | Versus ECG (bpm): Absolute bias = $-0.7 \pm 0.8$; Bias = $-0.3 \pm 0.1$; Intraclass correlation = 0.99 | <sup>121</sup> External, n=53 |
| | Ava bracelet | PPG sensor | Versus Polar chest strap, two electrode ECG (bpm): Bias = $7.8 \pm 3.9$; Pearson's $r = 0.92$, $p = 0.00$; Mean absolute percentage error = 11.4% | <sup>134</sup> External, n=37 |
| | Garmin Forerunner 245 | PPG sensor during a 3-min period as close as practicable to the start of each sleep period, i.e., ~30–60 min prior to lights out<sup>121</sup> | Versus ECG (bpm): Absolute bias = $5.4 \pm 12.6$; Bias = $5.0 \pm 12.8$; Intraclass correlation = 0.41 | <sup>121</sup> External, n=53 |
|  | Garmin Forerunner 45 | PPG sensor | Versus ECG (bpm): Bias = 0.67 at rest. During two sequential ramps from rest to exercise, bias was 10.82 bpm and 3.49 bpm. At steady state, exercise bias was 0.2 bpm. No difference with skin tone except during the second ramp (underestimated, >71% had <-5bpm difference) | <sup>126</sup> External, n=33 |
| | Apple Watch S6 | PPG sensor during sleep<sup>121</sup> | Versus ECG (bpm): Absolute bias = $1.5 \pm 1.5$; Bias = $0.5 \pm 2.1$; Intraclass correlation = 0.96 | <sup>121</sup> External, n=53 |
|  | Fitbit Charge 2 | PPG sensor during rest | Versus ECG (bpm): Non-significant difference=0.09, p=0.43. Bias = 0.51 bpm when heart rate < 50 bpm; Bias = -0.63 bpm when heart rate >80 bpm | <sup>155</sup> External, n=17 |
|  | Huawei Band 5 | PPG sensor | Limited published accuracy. Heart rate versus Polar chest strap, two electrode ECG (bpm) during elliptical exercise: r = 0.981 | <sup>156</sup> External, Huawei provided devices, n=10 |
| | | | Heart rate error range $\pm 10$ bpm | <sup>157</sup> Company site |
| Respiratory rate | Oura Ring 2.0 | Calculates rate via increase/decrease in heart rate with breath in/out | Versus ECG-calculation (respiratory rate in breaths/min): Correlation = 0.96; Mean Absolute Error = 0.71 | <sup>158</sup> Internal, n=43 |
|  | WHOOP 2.0 | Calculates median rate during sleep via increase/decrease in heart rate with breath in/out<sup>159</sup> (company site) | Versus respiratory inductive plethysmography (respiratory rate in breaths/min): Bias = 0.1; Percent Bias = 1.8%; Precision = 1.0; Precision Percent = 6.7% | <sup>129</sup> External, n=32 (21 female) |
|  | Garmin Forerunner 245 | Calculates rate via increase/decrease in heart rate with breath in/out | n/a | n/a |
|  | Ava bracelet | Calculates rate via increase/decrease in heart rate with breath in/out | Versus chest strap (respiratory rate in breaths/min): Mean Absolute Error = 0.9; Mean Absolute Percentage Error = 6.7% | <sup>160</sup> External, n= 7 |
|  | Apple Watch | Accelerometer-based estimate of nocturnal breaths per minute | n/a | n/a |
|  | Fitbit (Charge 2-4, Versa 1-2, Inspire HR) | Calculates rate via increase/decrease in heart rate with breath in/out<sup>161</sup> | Versus polysomnography (respiratory rate in breaths/min): Root Mean Squared Error = 0.648; Mean Absolute Error = 0.46; Mean Absolute Percentage Error = 3%; Bias = -0.24 | <sup>162</sup> Internal, n=28 |
| HRV | Oura Ring and Oura Ring 2.0 | PPG sensor every five minutes throughout the night - reports RMSSD | Versus ECG (RMSSD in ms): $r^2 = 0.980$; Bias = 1.2 | <sup>122</sup> Sponsored, n=49 |
| | | | Versus ECG (RMSSD in ms): $r^2 = 0.967$; Bias = -0.47, p<0.001; Range = 21 to 78 | <sup>123</sup> In-house, n=10 |
| | | | Versus ECG (RMSSD in ms): Absolute accuracy = $18.9 \pm 35.9$; Bias = $-10.2 \pm 39.4$; Intraclass correlation = 0.63 | <sup>121</sup> External, n=53 |
| | | | Versus ECG: RMSSD $r^2$ values > 0.9; Alternative HRV measures: $r^2$ values of SDNN (0.5), AVNN (0.8), pNN50 (0.8), LF (0.4), and HF (0.6) variable, all $p < 0.001$ | <sup>163</sup> External, n=35 |
| | WHOOP 3.0 | PPG sensor in non-wake in primary sleep episode every 30 seconds - reports RMSSD<sup>154</sup> | Versus ECG (RMSSD in ms): Absolute bias = $4.7 \pm 3.6$; Bias = $-4.5 \pm 3.9$; Intraclass correlation = 0.99 | <sup>121</sup> External, n=53 |
| | Ava bracelet | PPG sensor<sup>164</sup> (company site) - reports LF/HF ratio | Versus actigraph with Polar chest strap, 2 electrode ECG (LF/HF ratio): $r = -0.28$, $p = 0.00$ | <sup>134</sup> External, n=33 |
| | Garmin Forerunner 245 | PPG sensor - reports SDRR and RMSSD | Versus ECG (RMSSD): Absolute bias = $33.1 \pm 39.9$; Bias = $-22.4 \pm 46.9$; Intraclass correlation = 0.24 | <sup>121</sup> External, n=53 |
| | Apple Watch S6 | PPG sensor - reports RMSSD | Versus ECG (RMSSD in ms): Absolute bias = $22.5 \pm 19.2$; Bias = $-9.6 \pm 28.1$; Intraclass correlation = 0.67 | <sup>121</sup> External, n=53 |
|  | Fitbit Sense | PPG sensor - reports RMSSD | n/a | n/a |
|  | Huawei Band 5 | PPG sensor - type of HRV measurement unknown | n/a | n/a |
| Sleep | Oura Ring 2.0 | Accelerometer + temperature sensor + PPG sensor for 4-stage categorization (wake, light, deep, REM) | Versus polysomnography (sleep duration in mins): Accuracy = $0.88 \pm 0.05$; Sensitivity = $0.93 \pm 0.05$; Specificity = $0.49 \pm 0.22$ | <sup>165</sup> External, n=29 |
| | | | Versus polysomnography: (sleep vs wake) Accuracy = 89%; (sleep duration in mins) Bias = $1.5 \pm 40.9$ | <sup>121</sup> External, n=53 |
| | WHOOP 3.0 or 4.0 | PPG sensor + accelerometer for 4-stage categorization (wake, light, deep, REM) | Versus polysomnography: (sleep vs wake) Accuracy = 86%; (sleep duration in mins) Bias = $-12.2 \pm 36.3$ | <sup>121</sup> External, n=53 |
| | Garmin Forerunner 245 | Accelerometer + PPG sensor + SpO2 sensor for 4-stage categorization (wake, light, deep, REM) | Versus polysomnography: (sleep vs wake) Accuracy = 89%; (sleep duration in mins) Bias = $43.8 \pm 38.0$ | <sup>121</sup> External, n=53 |
|  | Ava bracelet | Accelerometer | Versus actigraphy: (sleep duration in hours)<br>Mean Absolute Percent Error = 8.5%; Pearson's r = 0.73, p=0.00; Bias = 0.18 ± 0.88 | <sup>134</sup> External, n=33 |
|  | Apple Watch S6 | PPG sensor in 5 min epochs; 3-stage categorization (wake, light, deep) | Versus polysomnography: (sleep vs wake) Accuracy = 88%; (sleep duration in mins) Bias = 39.5 ± 41.5 | <sup>121</sup> External, n=53 |
|  | Fitbit (Alta<sup>133</sup>, Charge<sup>165</sup>, Non-sleep-staging models = Charge HR, Flex, Alta, One, Ultra, Classic; Sleep-staging models = Surge, Alta HR, Charge 2, Versa) | PPG sensor + accelerometer for 4-stage categorization (wake, light, deep, REM)<sup>166</sup> | Versus actigraphy: (sleep duration in minutes)<br>Intraclass correlation = 0.84; Mean Absolute Error = 56 | <sup>133</sup> External, n=30 |
|  |  |  | Versus polysomnography: (sleep duration in minutes)<br>Accuracy = 0.84 ± 0.07; Sensitivity = 0.89 ± 0.09; Specificity = 0.49 ± 0.18 | <sup>165</sup> External, n=27 |
|  |  |  | *Older sleep-staging models using movement only:*<br>Versus polysomnography: (sleep duration in minutes) Bias = 7-67 mins; (wake duration in minutes) Bias = 6-44; (sleep efficiency) Bias = 2-15%; No difference in sleep onset latency. Sensitivity = 0.87-0.99; Specificity = 0.1-0.52<br>*Newer sleep-staging models using HRV and movement:*<br>Versus polysomnography: Newer models show no difference for sleep duration, wake after sleep onset, but underestimated sleep onset latency. Sensitivity = 0.95-0.96; Specificity = 0.58-0.69 | <sup>167</sup> Systematic review with 8 papers with quantitative data, n=7 to 63 |
|  | Huawei Band GT | Uses accelerometry, HR, HRV for 4-stage categorization (wake, light, deep, REM)<sup>168</sup> (company site) | Versus actigraphy (sleep time assessment): Intraclass correlation: 0.25; Mean Absolute Percentage Error = 23.29% | <sup>169</sup> External, n=102 |

PPG= photoplethysmography, ECG = electrocardiogram, RMSSD = root mean square of successive differences, SDNN = Standard Deviation of Normal-to-Normal Intervals, AVNN=Average of the Normal-to-Normal (NN) intervals, pNN50 = percentage of Normal-to-Normal (NN) intervals that differ by more than 50 milliseconds from the preceding interval, LF = low frequency, HF=high frequency, SDRR = standard deviation between consecutive R-wave

#### 2.4.1 Skin temperature

Many wearables now passively monitor skin temperature using negative temperature coefficient sensors that contain semiconductive materials that decrease their resistance with increasing temperature. Lab-based studies show a body temperature change of 0.3-0.7°C across the menstrual cycle^21,77^ and 1.1°C during perimenopausal hot flushes, based on the gold-standard rectal thermometer^114,115^. While reported accuracy compared to core body temperature across all wearables is lacking (Table 3), an in-house study of the Oura Ring demonstrated a high correlation (r² >0.92), including an accuracy of 0.36°C and precision of 0.13°C when compared to skin temperature measured with i-Button monitors^116^. The i-Button is a research-grade wireless semiconductor skin-temperature sensor^117^ also used as a standard in some validation studies. Comparison of skin temperature immediately after wake measured with an Oura Ring versus oral temperature showed a correlation of r=0.56^79^. These results suggest that the Oura Ring is likely sufficiently accurate to capture changes in temperature across the menstrual cycle and in response to hot flushes. But researchers must keep in mind that skin temperature is known to systematically present around 2°C lower than^118^, and out of phase^119^ with core body temperature changes. BMI can also impact the absolute value of recordings due to insulation with visceral fat, although magnitude of change is unaffected^80^.

#### 2.4.2 Resting heart rate

Wearables use photoplethysmography sensors that detect minor variations in the intensity of light transmitted through the skin to estimate changes in blood flow and cardiac activity. Studies using gold-standard electrocardiograms (ECGs) have shown that resting heart rate increases by 2.3 to 3 bpm between the follicular and the luteal phase of the menstrual cycle^88,120^ while changes during menopause are unknown. In one externally conducted study, six devices (Apple Watch S6, Garmin Forerunner 245 Music, Polar Vantage V, Oura Ring 2.0, WHOOP 3.0, and Somfit) were tested overnight in 53 participants. Compared to ECG, the WHOOP 3.0 device demonstrated the highest intraclass correlation of 0.99 for nocturnal heart rate and lowest average over-estimation bias of 0.7±0.8 bpm^121^. Most devices with nocturnal recordings available reported a good intraclass correlation above 0.85 and low bias of less than 2.6 bpm (Table 3), with the exception being a 0.65 intraclass correlation reported with Somfit, which is a head-worn sensor and was not used in any of the studies we reported. The Oura Ring 2.0 showed an intraclass correlation of 0.85 and overestimation bias of 1.8bpm, but an Oura-sponsored^122^ and an in-house study^123^ reported higher r^2^ values of 0.99 and smaller mean biases of −0.63.

Together, these results suggest that many of the commercially available ring and wrist-worn devices are sufficiently accurate to measure changes in resting heart rate across the menstrual cycle, but there are considerations to keep in mind. For example, discrepancies in data resolution and sampling windows (e.g., raw versus summary data) can hinder comparison between devices. Specifically, the Garmin devices provided pre-sleep instead of nocturnal cardiac data, which may have contributed to reported lower accuracy as it may not have captured a true resting heart rate^121^.

There is some evidence that photoplethysmography-measured heart rate can be influenced by BMI^124^ and skin tone^125^, but the temporal location of the peaks used to detect heart rate remained stable. A study using Garmin devices during rest and activity found that ECG- and PPG-measured heart rates only significantly differed during a transition from rest to activity^126^. Taken together, the evidence to-date indicates that measures of resting heart rate may be fairly robust across these demographic characteristics with scope for further validation and improvement.

#### 2.4.3 Respiratory rate

Wearable devices estimate respiratory rate using small photoplethysmography sensors on the skin that detect the high-frequency variations in heart rate linked to respiratory cycles, a feature called respiratory sinus arrhythmia. In the laboratory, respiratory rate during sleep is measured with belts that sense chest wall motion or with respiratory inductive plethysmography where coils of wire are placed around the chest and abdomen^127^. Studies using this approach show that respiratory rate changes across the menstrual cycle range from 0.63 to 2 breaths/min^97,128^. We found limited data on wearable accuracy for respiratory rate (Table 3). One external study funded by a grant from WHOOP reported no significant difference between recordings with WHOOP 2.0 and respiratory inductive plethysmography (15.7±1.7 versus 15.6±1.7 breaths/min, respectively), with a mean bias of 0.1 breaths/min (1.8%) and a precision bias of 1.0 breaths/min (6.7%)^129^, providing some confidence in using photoplethysmography on a wrist-worn device to measure changes in respiratory rate related to the menstrual cycle, with additional validation needed for larger and diverse cohorts and for other wearables.

#### 2.4.4 Heart rate variability (HRV)

Wearables estimate HRV using heart rate recordings from the photoplethysmography sensor. In-lab studies use electrocardiograms to record individual heart beats then calculate the variability in heart beat timing, most commonly using root mean square of successive differences (RMSSD) between the interbeat intervals over a sleep or rest period, reported in milliseconds or log-transformed values. A meta-analysis of 37 in-lab studies indicated a significant overall decrease in cardiac vagal activity from the follicular to the luteal phase, but only 2 of 8 studies reported a significant difference in RMSSD in the range of 9-20 ms^98,130,131^.

The comparative study of Miller et al. found that the WHOOP 3.0 device demonstrated the highest HRV estimation accuracy compared to ECG measures, with a bias of −4.5±3.9 ms with a 0.99 intraclass correlation^121^. An external study funded by an Oura grant reported a 0.98 r^2^ value and −1.2ms bias when compared to ECG measurements^122^, which is an improved performance from the 0.85 intraclass correlation reported in the six device comparative study^121^. These results indicate researchers should be aware of underestimation bias according to their device, as underestimation could be up to 50% of the variability seen across the menstrual cycle.

#### 2.4.5 Sleep

Detection of sleep using commercial wearable devices typically uses a combination of cardiac metrics such as resting heart rate, HRV, respiratory rate, and movement metrics in proprietary sleep-detection algorithms, while polysomnography is the gold standard for measuring sleep in a laboratory^127^. The comparative study of six wearables found that all could perform the binary classification of sleep vs. wake periods throughout the night with relatively high accuracy (above 85%) compared to polysomnography. However, the accuracy in estimating total sleep duration varied widely, with mean errors on the order of one minute overestimation for the Oura Ring and Polar watch, and 40 minutes or more for the Apple Watch and Garmin watch. All six devices showed low accuracy in identifying the sleep stages (e.g., slow-wave sleep versus rapid eye movement (REM) sleep), reporting less than 65% agreement with polysomnography^121^. Several other external studies show that commercial wearable devices tend to overestimate sleep duration. Sleep duration was overestimated by 15 minutes with the Oura Ring^132^, by 56 minutes with the Fitbit^133^, and 11 minutes with the Ava bracelet)^134^. Taken together, sleep stage estimation is inaccurate across ring and wrist-worn devices, while researchers prioritising estimated total sleep accuracy should be aware of the variation in accuracy across devices and choose devices accordingly.

## 3. Discussion

This scoping review reveals several promising contributions of digital health studies to understanding of the interplay between physiology, behaviours, symptoms, and hormonally-driven processes including the menstrual cycle and menopausal transition. Studies using digital health tools have begun to establish detailed, normative biometric data across the menstrual cycle. For example, while skin temperature is often used to estimate fertile windows, we found that digital health studies indicate that the largest decrease in skin temperature, and in some cases, the minimum temperature can occur as early as day five of the menstrual cycle, unlike the 1-2 days before ovulation reported by lab studies. This knowledge could improve the efficacy of wearable-based tools for contraception or conception. Digital health studies have also provided additional evidence to support fluctuations in resting heart rate and HRV across the menstrual cycle, and discovered a decrease in the magnitude of this fluctuation with age.

Understanding these changes across the lifespan can help women set realistic training and recovery expectations. In addition, digital health tools have begun to address how biometric changes are connected to hormone-related symptoms. For instance, studies using data from wearables found an inverse association with HRV and symptoms (both affective and physiological) in women with self-reported premenstrual disorder symptoms. This information is needed for quantifying the effectiveness of symptom-reducing interventions.

Despite new data from digital health studies to characterise normative trends, our review of studies to-date reveals a need to better understand deviations from the norm, both within and between individuals. In some cases, whole cohorts have been omitted from most research. For example, most of the research we reviewed was carried out in predominantly White cohorts and in the United States, United Kingdom, and Scandinavian countries, which limits our understanding of the relationships between race, ethnicity, and menstrual cycle physiology.

Perimenopausal women were often omitted from studies, so characterisation of normal hormonal fluctuations and physiological change during perimenopause is lacking. This is reflected by the fact that fewer than 1 in 10 women in the United Kingdom feel that they have received sufficient advice on what to expect during perimenopause^135^, with similar trends in the United States, where 64% of adult women reported a need for more doctors specialising in menopause^136^. Furthermore, we found that many studies focused on regularly menstruating women, often only including analysis of cycles with estimated ovulation, and excluding women with conditions impacting menstruation or using hormonal contraceptives. Menstrual conditions are common; endometriosis affects 10% of women and many women are undiagnosed for years^137,138^. Although some atypical menstrual cycles may present no health concerns, identifying and understanding the causes of irregularity and anovulation and their biometric connections becomes crucial for better diagnostics and monitoring intervention efficacy. The relative accessibility and lower cost of digital health tools compared to lab-based approaches offers promise to including these under-studied cohorts in research.

Future digital health studies should concurrently measure hormones, biometrics, and menstrual-associated symptoms. While we expected to find many studies exploring relationships between these measures, studies to-date were few. For example, the physiological impact of hormonal contraception, including effects on cardiovascular biometrics associated with athletic performance remains under-characterised. For female athletes striving for peak performance, even small physiological changes may be significant, yet women are often left to use trial and error approaches when selecting contraception. Furthermore, we need to understand the timing and characteristics of the transition to a natural hormone cycle after individuals discontinue using hormonal contraception^139^. Using wearables could help to characterise these effects and contribute to informed contraceptive choices, but studies have not yet explored these relationships. Further, the connections between symptoms, biometrics, and hormonal fluctuations in the menopausal transition is markedly understudied.

Digital health tools can help quantify how behaviour and physical or social environment may influence hormones and physiology, enabling the development of guidelines to manage symptoms and optimise performance, but again, we found few studies that have explored these areas. For example, future digital health studies could examine whether altering sleep in the luteal phase could mitigate premenstrual symptoms, or quantify the relationships between physical activity and the menstrual cycle, bringing much-needed evidence to guide the popular trend of “cycle syncing”. By understanding how behaviours such as sleep and exercise can mitigate or exacerbate symptoms related to the menstrual cycle or perimenopause, women could be empowered to manage their health and performance more effectively. Wearables could also serve as intervention tools. For instance, in a pilot study, a mobile app providing HRV biofeedback training showed promise in reducing premenstrual anxiety, stress, and poor sleep^140^.

We found that most commercial wearables used in the studies we reviewed had sufficient accuracy to capture changes in heart rate, respiratory rate, and to a lesser degree HRV. While sensors that measure skin temperature appear sufficiently accurate to capture changes across the menstrual cycle, this is based on only one study. Results for accuracy of sleep duration were device-dependent, and other sleep characteristics (like sleep staging), should be used with caution. The superior accuracy for cardiac biometrics likely reflects technological maturity of sensors and extensive validation history compared to more recently integrated features like sleep and skin temperature, suggesting accuracy will likely improve as hardware and signal processing algorithms evolve. The reported accuracies mirror the consistency of the findings in Section 2.2, highlighting how improvements in sensor technology combined with access to continuous monitoring in large cohorts enables higher resolution data and statistical power that can clarify mixed reports and reveal new insights. Additional research is still needed to validate accuracy of wearables in cohorts with diverse characteristics (e.g., BMI, skin color, and presence of tattoos), given the limited research to-date in this area. Furthermore, retrospective studies using passively collected wearable data may contain self-selection bias as individuals who purchase fitness tracking wearables may be more active, of a higher socioeconomic status, and more health conscious than average. However, providing wearable devices to participants and intentionally recruiting diverse populations can help to mitigate this bias.

Researchers selecting digital health tools must consider their cohort, research question, and the limitations of each tool. For instance, self-reports cannot confirm if a menstrual cycle is truly absent or due to a lapse in reporting of menses onset. While biometric data can be used to predict menstrual cycle and/or ovulation occurrence with good accuracy, ovulation can sometimes occur without the typical biometric pattern changes and vice versa. A study of endurance runners may require a device with long battery life that measures heart rate and distance with high accuracy and resolution during activity only, while a study focused on recovery may require capturing accurate nocturnal cardiac and sleep parameters with constant daily activity monitoring. As the wearables we reviewed are wrist or finger-worn, it is important to note that devices using 2-electrode chest straps^141^ and upper-arm-photoplethysmography sensors can be more accurate than wrist-worn devices^142^ for measuring heart rate during activity.

As technology continues to improve, we hope for the development of new tools to monitor hormones, physiology, and behaviour alongside consistent protocols and safe practices. Continuous and minimally invasive monitoring of biomarkers, such as hormones via sweat or interstitial fluid hold promise. Such monitoring could provide insights into normal, optimal, and pathological states, ultimately guiding more effective management of symptoms and health. As research in this field grows, it is important for researchers to establish and use standardised methodologies (e.g., measuring HRV during consistent sleep periods—such as the final hour before wake—versus varying times throughout the day) and data reporting (e.g., consistent HRV metrics). With the increasing use and development of digital health tools in women’s health research, there is also a vital need to address privacy concerns, particularly given the legal complexities surrounding pregnancy termination. Careful consideration must be given to research planning, data handling, and privacy, as discussed in detail elsewhere^143^, including specific considerations for female health data and privacy analyses of existing apps^144,145^.

Digital health research offers promise in improving our understanding of the interaction of physiology, behaviour, and symptoms during the menstrual cycle and perimenopause. The use of digital health tools in female health is often focused on pregnancy prevention and fertility. By facilitating characterisation of healthy menstrual cycle norms across a more diverse group of women, digital health tools can enable understanding of healthy norms and identify when medical evaluation is warranted. Furthermore, integration of biometric monitoring with underlying hormone fluctuations, symptoms, and health behaviours can help to identify targets to monitor interventions and develop healthy biometric ranges. Not only is this understanding a vital first step toward developing effective solutions, but it also empowers women to take ownership of their health and enhance their quality of life.

## 4. Methods

Our scoping review adhered to the recommendations outlined in the PRISMA guidelines (https://www.prisma-statement.org/scoping). We searched for articles on Google Scholar, Web of Science Core Collection, and PubMed through December 18, 2025 with the terms (“digital health” OR “wearable technology” OR “wearable devices” OR “mobile health” OR “smartphone application” OR “app”) AND (“scientific” OR “research outcomes” OR “insights” OR “study results” OR “clinical research”). We combined these digital health terms first with (“menstrual cycle” OR “menstruation” OR “period tracking”), and then with (“ovulation” OR “ovulatory cycle” OR “cycle tracking”).

We removed duplicates, then two authors (S.J. and J.O.) independently reviewed titles, abstracts, and keywords of all the retrieved articles. Any disagreements regarding study eligibility were resolved through discussion and consensus between the two reviewers. Articles were included if they (i) were written in English; (ii) involved human participants; (iii) used wearable devices or mobile health apps; (iv) analysed female health physiology or biometrics; and (v) provided quantitative data collected with digital health tools. We excluded dissertations, theses, conference proceedings, conference abstracts, and reviews. We excluded articles that did not include menstrual tracking with a digital health tool and/or articles where longitudinal data concurrent with menstrual tracking was not reported (Figure 1). We excluded reviews as primary articles, but reviewed reference lists to identify any additional papers that met our criteria. We repeated the process replacing the first grouping of search terms with “menopaus* OR perimenopaus*”.

We extracted the following outcomes from the included articles: scientific insights on female health with regard to menstruation and menopause; how menstrual cycle phase was determined (e.g., self report or hormonal testing); and study details including number of participants, menstrual cycles, participant demographics (age, BMI, ethnicity), and the digital health tool(s) used in the study.

## Contributions

SCJ, JO, and JH conceived and defined the scope of the review and wrote the manuscript. SCJ and JO carried out the scoping review search and interpreted, collated and processed the data. SCJ, JO, EK, SD, and JH reviewed, edited, and approved the final manuscript.

## Data Availability

Data sharing is not applicable to this article as no datasets were generated or analysed during the current study.

## Acknowledgements

This study was funded by the Wu Tsai Human Performance Alliance at Stanford University and the Joe and Clara Tsai Foundation, as well as the National Institutes of Health through Grant P41EB027060. The funders played no role in study design, data collection, analysis and interpretation of data, or the writing of this manuscript. We thank Dr Alex Gonzalez for his contribution to early conception.

## References

1. Tanaka, E. et al. Burden of menstrual symptoms in Japanese women: results from a survey-based study. J. Med. Econ. 16, 1255–1266 (2013).

2. Rapkin, A. J. & Winer, S. A. Premenstrual syndrome and premenstrual dysphoric disorder: quality of life and burden of illness. Expert Rev. Pharmacoecon. Outcomes Res. 9, 157–170 (2009).

3. Bruinvels, G. et al. Prevalence and frequency of menstrual cycle symptoms are associated with availability to train and compete: a study of 6812 exercising women recruited using the Strava exercise app. Br. J. Sports Med. 55, 438–443 (2021).

4. Halbreich, U., Borenstein, J., Pearlstein, T. & Kahn, L. S. The prevalence, impairment, impact, and burden of premenstrual dysphoric disorder (PMS/PMDD). Psychoneuroendocrinology 28 **Suppl 3**, 1–23 (2003).

5. Schoep, M. E. et al. Productivity loss due to menstruation-related symptoms: a nationwide cross-sectional survey among 32 748 women. BMJ Open 9, e026186; 10.1136/bmjopen-2018-026186 (2019).

6. Ponzo, S. et al. Menstrual cycle-associated symptoms and workplace productivity in US employees: A cross-sectional survey of users of the Flo mobile phone app. *Digit*. Health 8; 10.1177/20552076221145852 (2022).

7. Starr, M. et al. Epidemiology of menstrual-related absenteeism in 44 low-income and middle-income countries: a cross-sectional analysis of Multiple Indicator Cluster Surveys. Lancet Glob. Health 13, e285–e297 (2025).

8. Solli, G. S., Sandbakk, S. B., Noordhof, D. A., Ihalainen, J. K. & Sandbakk, Ø. Changes in self-reported physical fitness, performance, and side effects across the phases of the menstrual cycle among competitive endurance athletes. Int. J. Sports Physiol. Perform. 15, 1324–1333 (2020).

9. Armour, M., Parry, K. A., Steel, K. & Smith, C. A. Australian female athlete perceptions of the challenges associated with training and competing when menstrual symptoms are present. Int. J. Sports Sci. Coach. 15, 316–323 (2020).

10. Findlay, R. J., Macrae, E. H. R., Whyte, I. Y., Easton, C. & Forrest Née Whyte, L. J. How the menstrual cycle and menstruation affect sporting performance: experiences and perceptions of elite female rugby players. Br. J. Sports Med. 54, 1108–1113 (2020).

11. Badawy, Y., Spector, A., Li, Z. & Desai, R. The risk of depression in the menopausal stages: A systematic review and meta-analysis. J. Affect. Disord. 357, 126–133 (2024).

12. Coronado, P. J. et al. Quality of life at peri- and postmenopause: analysis of the results from app ‘Mi Menopausia’. Maturitas 197, 108267 (2025).

13. Faubion, S. S. et al. Impact of Menopause Symptoms on Women in the Workplace. Mayo Clin. Proc. 98, 833–845 (2023).

14. Rodriguez, L. A., Casey, E., Crossley, E., Williams, N. & Dhaher, Y. Y. The hormonal profile in women using combined monophasic oral contraceptive pills varies across the pill cycle: a temporal analysis of serum endogenous and exogenous hormones using liquid chromatography with tandem mass spectroscopy. Am. J. Physiol. Endocrinol. Metab. 327, E121–E133 (2024).

15. Stanford, J. B. & Mikolajczyk, R. T. Mechanisms of action of intrauterine devices: update and estimation of postfertilization effects. Am. J. Obstet. Gynecol. 187, 1699–1708 (2002).

16. Burger, H. The menopausal transition--endocrinology. J. Sex. Med. 5, 2266–2273 (2008).

17. Hale, G. E., Robertson, D. M. & Burger, H. G. The perimenopausal woman: endocrinology and management. J. Steroid Biochem. Mol. Biol. 142, 121–131 (2014).

18. Forman, R. G., Chapman, M. C. & Steptoe, P. C. The effect of endogenous progesterone on basal body temperature in stimulated ovarian cycles. Hum. Reprod. 2, 631–634 (1987).

19. Israel, L. S. & Schneller, O. The thermogenic property of progesterone. Obstet. Gynecol. Surv. 5, 532–533 (1950).

20. Stachenfeld, N. S., Silva, C. & Keefe, D. L. Estrogen modifies the temperature effects of progesterone. J. Appl. Physiol. (1985) 88, 1643–1649 (2000).

21. Baker, F. C., Siboza, F. & Fuller, A. Temperature regulation in women: Effects of the menstrual cycle. Temperature (Austin) 7, 226–262 (2020).

22. Behan, M. & Wenninger, J. M. Sex steroidal hormones and respiratory control. Respir. Physiol. Neurobiol. 164, 213–221 (2008).

23. Haggerty, C. L., Ness, R. B., Kelsey, S. & Waterer, G. W. The impact of estrogen and progesterone on asthma. Ann. Allergy Asthma Immunol. 90, 284–91; quiz 291–3, 347 (2003).

24. Blaser, B. L., Weymar, M. & Wendt, J. Premenstrual syndrome is associated with differences in heart rate variability and attentional control throughout the menstrual cycle: A pilot study. Int. J. Psychophysiol. 204, 112374 (2024).

25. Matsumoto, T., Ushiroyama, T., Kimura, T., Hayashi, T. & Moritani, T. Altered autonomic nervous system activity as a potential etiological factor of premenstrual syndrome and premenstrual dysphoric disorder. Biopsychosoc. Med. 1, 24 (2007).

26. Baker, F. C., Colrain, I. M. & Trinder, J. Reduced parasympathetic activity during sleep in the symptomatic phase of severe premenstrual syndrome. J. Psychosom. Res. 65, 13–22 (2008).

27. Landén, M. et al. Heart rate variability in premenstrual dysphoric disorder. Psychoneuroendocrinology 29, 733–740 (2004).

28. Pattnaik, S., Das, D. & Venkatesan, V. A. Validation of urinary reproductive hormone measurements using a novel smartphone connected reader. Sci. Rep. 13, 9227; 10.1038/s41598-023-36539-w (2023).

29. Pichon, A., Jackman, K. B., Winkler, I. T., Bobel, C. & Elhadad, N. The messiness of the menstruator: assessing personas and functionalities of menstrual tracking apps. J. Am. Med. Inform. Assoc. 29, 385–399 (2022).

30. Zvarikova, K., Machova, V. & Pera, A. Menstrual cycle tracking apps, fertility and reproductive data, and mobile health care management. J. Res. Gend. Stud. 12, 84 (2022).

31. Schantz, J. S., Fernandez, C. S. P. & Anne Marie, Z. J. Menstrual cycle tracking applications and the potential for epidemiological research: a comprehensive review of the literature. Curr. Epidemiol. Rep. 8, 9–19 (2021).

32. Lyzwinski, L., Elgendi, M. & Menon, C. Innovative approaches to menstruation and fertility tracking using wearable reproductive health technology: systematic review. J. Med. Internet Res. 26, e45139; 10.2196/45139 (2024).

33. Kalampalikis, A., Chatziioannou, S. S., Protopapas, A., Gerakini, A. M. & Michala, L. mHealth and its application in menstrual related issues: a systematic review. Eur. J. Contracept. Reprod. Health Care 27, 53–60; 10.1080/13625187.2021.1980873 (2022).

34. Saugar, E. E. et al. Factors used by mobile applications to predict female fertility status and their reported effectiveness: A scoping review. Cureus 15, e48847; 10.7759/cureus.48847 (2023).

35. Rabe, I. & Ehlers, J. P. Reliability of Cycle Applications for Pregnancy Planning and Contraception: A Systematic Review. Mayo Clin. Proc. Digit. Health 3, 100239; 10.1016/j.mcpdig.2025.100239 (2025).

36. Huhn, S. et al. The impact of wearable technologies in health research: scoping review. JMIR Mhealth Uhealth 10, e34384; 10.2196/34384 (2022).

37. Fitzpatrick, M. B. & Thakor, A. S. Advances in Precision Health and Emerging Diagnostics for Women. J. Clin. Med. Res. 8, (2019).

38. Moghimikandelousi, S. et al. Advances in biomonitoring technologies for women’s health. Nature Communications 16, 8507 (2025).

39. Rosen Vollmar, A. K., Mahalingaiah, S. & Jukic, A. M. The menstrual cycle as a vital sign: a comprehensive review. F S Rev. 6, 100081 (2025).

40. Committee on Practice Bulletins—Gynecology. Practice bulletin no. 128: diagnosis of abnormal uterine bleeding in reproductive-aged women. Obstet. Gynecol. 120, 197–206 (2012).

41. Fraser, I. S., Critchley, H. O. D., Broder, M. & Munro, M. G. The FIGO recommendations on terminologies and definitions for normal and abnormal uterine bleeding. Semin. Reprod. Med. 29, 383–390 (2011)

42. Nguyen, B. T. et al. Detecting variations in ovulation and menstruation during the COVID-19 pandemic, using real-world mobile app data. PLoS One 16, e0258314; 10.1371/journal.pone.0258314 (2021).

43. Soumpasis, I., Grace, B. & Johnson, S. Real-life insights on menstrual cycles and ovulation using big data. Hum. Reprod. Open 2020, hoaa011 (2020).

44. Grieger, J. A. & Norman, R. J. Menstrual cycle length and patterns in a global cohort of women using a mobile phone app: retrospective cohort study. J. Med. Internet Res. 22, e17109; 10.2196/17109 (2020).

45. Bull, J. R. et al. Real-world menstrual cycle characteristics of more than 600,000 menstrual cycles. NPJ Digit. Med. 2, 83; 10.1038/s41746-019-0152-7 (2019).

46. Symul, L., Wac, K., Hillard, P. & Salathé, M. Assessment of menstrual health status and evolution through mobile apps for fertility awareness. NPJ Digit. Med. 2, 64; 10.1038/s41746-019-0139-4 (2019).

47. Sohda, S., Suzuki, K. & Igari, I. Relationship between the menstrual cycle and timing of ovulation revealed by new protocols: analysis of data from a self-tracking health app. J. Med. Internet Res. 19, e391; 10.2196/jmir.7468 (2017).

48. Markovic, A. et al. Physiological response to the COVID-19 vaccine: insights from a prospective, randomized, single-blinded, crossover trial. J. Med. Internet Res. 26, e51120; 10.2196/51120 (2024).

49. Faust, L. et al. Findings from a mobile application-based cohort are consistent with established knowledge of the menstrual cycle, fertile window, and conception. Fertil. Steril. 112, 450–457.e3 (2019).

50. Thigpen, N., Patel, S. & Zhang, X. Oura Ring as a tool for ovulation detection: validation analysis. J. Med. Internet Res. 27, e60667; 10.2196/60667 (2025).

51. Li, K. et al. Characterizing physiological and symptomatic variation in menstrual cycles using self-tracked mobile-health data. NPJ Digit. Med. 3, 79; 10.1038/s41746-020-0269-8 (2020).

52. Jasinski, S. R., Presby, D. M., Grosicki, G. J., Capodilupo, E. R. & Lee, V. H. A novel method for quantifying fluctuations in wearable derived daily cardiovascular parameters across the menstrual cycle. NPJ Digit. Med. 7, 373; 10.1038/s41746-024-01394-0 (2024).

53. Cunningham, A. C. et al. Chronicling menstrual cycle patterns across the reproductive lifespan with real-world data. Sci. Rep. 14, 10172; 10.1038/s41598-024-60373-3 (2024).

54. Roffler, A., Fleddermann, M.-T., de Haan, H., Krüger, K. & Zentgraf, K. Menstrual cycle tracking in professional volleyball athletes. Front. Sports Act. Living 6, 1408711; 10.3389/fspor.2024.1408711 (2024).

55. Bradley, D. et al. Time to conception and the menstrual cycle: an observational study of fertility app users who conceived. Hum. Fertil. (Camb) 24, 267–275 (2021).

56. Ramaiyer, M. et al. The association of COVID-19 vaccination and menstrual health: A period-tracking app-based cohort study. Vaccine X 19, 100501 (2024).

57. Alvergne, A., Vlajic Wheeler, M. & Högqvist Tabor, V. Do sexually transmitted infections exacerbate negative premenstrual symptoms? Insights from digital health. Evol. Med. Public Health 2018, 138–150 (2018).

58. Vieira, C. S. et al. Real-world validation of a bleeding prediction algorithm in levonorgestrel intrauterine device users using the MyIUS mobile app. Contraception 152, 111201 (2025).

59. Frenz, A.-K., Ahlers, C., Beckert, V., Gerlinger, C. & Friede, T. Predicting menstrual bleeding patterns with levonorgestrel-releasing intrauterine systems. Eur. J. Contracept. Reprod. Health Care 26, 48–57 (2021).

60. Beckert, V. et al. Bleeding patterns with the 19.5 mg LNG-IUS, with special focus on the first year of use: implications for counselling. Eur. J. Contracept. Reprod. Health Care 24, 251–259 (2019).

61. The Practice Committee of the American Society for Reproductive Medicine. The clinical relevance of luteal phase deficiency: a committee opinion. Fertil. Steril. 98, 112–1117.

62. Crawford, N. M., Pritchard, D. A., Herring, A. H. & Steiner, A. Z. Prospective evaluation of luteal phase length and natural fertility. Fertil. Steril. 107, 749–755 (2017).

63. De Souza, M. J. et al. High prevalence of subtle and severe menstrual disturbances in exercising women: confirmation using daily hormone measures. Hum. Reprod. 25, 491–503 (2010).

64. Beckmann, C. R. B. et al. Preconception and Antepartum Care in Obstetrics and Gynecology, 7th ed. (eds. Beckman, C. R. B., Herbert, W., Laube, D., Ling, F. & Smith, R) 61–78 (Lippincott Williams & Wilkins, Philadelphia, PA, 2013).

65. Lenton, E. A., Landgren, B. M., Sexton, L. & Harper, R. Normal variation in the length of the follicular phase of the menstrual cycle: effect of chronological age. Br. J. Obstet. Gynaecol. 91, 681–684 (1984).

66. Wegrzynowicz, A. K. et al. Complete cycle mapping using a quantitative at-home hormone monitoring system in prediction of fertile days, confirmation of ovulation, and screening for ovulation issues preventing conception. Medicina (Kaunas) 58, (2022).

67. Baird, D. D. et al. Application of a method for estimating day of ovulation using urinary estrogen and progesterone metabolites. Epidemiology 6, 547–550 (1995).

68. Chiazze, L., Jr, Brayer, F. T., Macisco, J. J., Jr, Parker, M. P. & Duffy, B. J. The length and variability of the human menstrual cycle. JAMA 203, 377–380 (1968).

69. Li, H. et al. Menstrual cycle length variation by demographic characteristics from the Apple Women’s Health Study. NPJ Digit. Med. 6, 100; 10.1038/s41746-023-00848-1 (2023).

70. Harley, K. G., Watson, A., Robertson, S., Vitzthum, V. J. & Shea, A. Menstrual cycle characteristics of U.S. adolescents according to gynecologic age and age at menarche. J. Pediatr. Adolesc. Gynecol. 37, 419–425 (2024).

71. Roberts, R. E., Farahani, L., Webber, L. & Jayasena, C. Current understanding of hypothalamic amenorrhoea. Ther. Adv. Endocrinol. Metab. 11, 2042018820945854 (2020).

72. Hartz, A. J., Barboriak, P. N., Wong, A., Katayama, K. P. & Rimm, A. A. The association of obesity with infertility and related menstrual abnormalities in women. Int. J. Obes. 3, 57–73 (1979).

73. Yeung, E. H. et al. Adiposity and sex hormones across the menstrual cycle: the BioCycle Study. Int. J. Obes. (Lond) 37, 237–243 (2013).

74. Itoi, S. et al. Body mass index and menstrual irregularity in a prospective cohort study of smartphone application users. NPJ Womens Health 3; 10.1038/s44294-025-00065-z (2025).

75. Zhang, C. Y. et al. Abnormal uterine bleeding patterns determined through menstrual tracking among participants in the Apple Women’s Health Study. Am. J. Obstet. Gynecol. 228, 213.e1–213.e22 (2023).

76. Hurst, B. S., Davies, K., Milnes, R. C., Knowles, T. G. & Pirrie. A novel technique for confirmation of the day of ovulation and prediction of ovulation in subsequent cycles using a skin-worn sensor in a population with ovulatory dysfunction: a side-by-side comparison with existing basal body temperature algorithm and vaginal core body temperature algorithm. Front. Bioeng. Biotechnol. 10, 807139; 10.3389/fbioe.2022.807139 (2022).

77. Marshall, J. A field trial of the basal-body-temperature method of regulating births. Lancet 2, 8–10 (1968).

78. Martinez, A. R. et al. The reliability, acceptability and applications of basal body temperature (BBT) records in the diagnosis and treatment of infertility. Eur. J. Obstet. Gynecol. Reprod. Biol. 47, 121–127 (1992).

79. Maijala, A., Kinnunen, H., Koskimäki, H., Jämsä, T. & Kangas, M. Nocturnal finger skin temperature in menstrual cycle tracking: ambulatory pilot study using a wearable Oura ring. BMC Womens Health 19, 150; 10.1186/s12905-019-0844-9 (2019).

80. Shilaih, M. et al. Modern fertility awareness methods: wrist wearables capture the changes in temperature associated with the menstrual cycle. Biosci. Rep. 38; 10.1042/BSR20171279 (2018).

81. Zhu, T. Y. et al. The accuracy of wrist skin temperature in detecting ovulation compared to basal body temperature: prospective comparative diagnostic accuracy study. J. Med. Internet Res. 23, e20710; 10.2196/20710 (2021).

82. Murayama, Y., Tang, Z., Saito, K., Nakano, T. & Mitsui, M. Monitoring basal body temperature rhythm: Abdominal skin temperature measurements taken during sleep as simpler alternative. IEEE Access 12, 182374–182385; 10.1109/ACCESS.2024.3511341 (2024).

83. Lin, G. et al. Understanding wrist skin temperature changes to hormone variations across the menstrual cycle. NPJ Womens Health 2, 35; 10.1038/s44294-024-00037-9 (2024).

84. Alzueta, E. et al. Tracking sleep, temperature, heart rate, and daily symptoms across the menstrual cycle with the Oura Ring in healthy women. Int. J. Womens Health 14, 491–503; 10.2147/IJWH.S341917 (2022).

85. Goodale, B. M. et al. Wearable sensors reveal menses-driven changes in physiology and enable prediction of the fertile window: observational study. J. Med. Internet Res. 21, e13404; 10.2196/13404 (2019).

86. Su, H.-W., Yi, Y.-C., Wei, T.-Y., Chang, T.-C. & Cheng, C.-M. Detection of ovulation, a review of currently available methods. Bioeng. Transl. Med. 2, 238–246 (2017).

87. McKinley, P. S. et al. The impact of menstrual cycle phase on cardiac autonomic regulation. Psychophysiology 46, 904–911 (2009).

88. Baker, F. C. & Lee, K. A. Menstrual cycle effects on sleep. Sleep Med. Clin. 13, 283–294 (2018).

89. Sato, N., Miyake, S., Akatsu, J. & Kumashiro, M. Power spectral analysis of heart rate variability in healthy young women during the normal menstrual cycle. Psychosom. Med. 57, 331–335 (1995).

90. Leicht, A. S., Hirning, D. A. & Allen, G. D. Heart rate variability and endogenous sex hormones during the menstrual cycle in young women. Exp. Physiol. 88, 441–446 (2003).

91. Yu, J.-L. et al. Tracking of menstrual cycles and prediction of the fertile window via measurements of basal body temperature and heart rate as well as machine-learning algorithms. Reprod. Biol. Endocrinol. 20, 118 (2022).

92. Shilaih, M., de Clerck, V., Falco, L., Kübler, F. & Leeners, B. Pulse rate measurement during sleep using wearable sensors, and its correlation with the menstrual cycle phases, a prospective observational study. Sci. Rep. 7, 1294; 10.1038/s41598-017-01433-9 (2017).

93. Lang, A.-L. et al. Feasibility study on menstrual cycles with Fitbit device (FEMFIT): prospective observational cohort study. JMIR Mhealth Uhealth 12, e50135; 10.2196/50135 (2024).

94. Altini, M. & Plews, D. What is behind changes in resting heart rate and heart rate variability? A large-scale analysis of longitudinal measurements acquired in free-living. Sensors (Basel) 21; 10.3390/s21237932 (2021).

95. Sims, S. T., Ware, L. & Capodilupo, E. R. Patterns of endogenous and exogenous ovarian hormone modulation on recovery metrics across the menstrual cycle. BMJ Open Sport Exerc. Med. 7, e001047; 10.1136/bmjsem-2021-001047 (2021).

96. Pierson, E., Althoff, T., Thomas, D., Hillard, P. & Leskovec, J. Daily, weekly, seasonal and menstrual cycles in women’s mood, behaviour and vital signs. *Nat*. Hum. Behav. 5, 716–725; 10.1038/s41562-020-01046-9 (2021).

97. Godbole, G., Joshi, A. R. & Vaidya, S. M. Effect of female sex hormones on cardiorespiratory parameters. J. Family Med. Prim. Care 5, 822–824 (2016).

98. Schmalenberger, K. M. et al. A systematic review and meta-analysis of within-person changes in cardiac vagal activity across the menstrual cycle: implications for female health and future studies. J. Clin. Med. 8; 10.3390/jcm8111946 (2019).

99. Ramesh, A., Nayak, T., Beestrum, M., Quer, G. & Pandit, J. A. Heart rate variability in psychiatric disorders: a systematic review. Neuropsychiatr. Dis. Treat. 19, 2217–2239 (2023).

100. Baker, F. C., Mitchell, D. & Driver, H. S. Oral contraceptives alter sleep and raise body temperature in young women. Pflügers Arch. 442, 729–737 (2001).

101. Martin, J. G. & Buono, M. J. Oral contraceptives elevate core temperature and heart rate during exercise in the heat. Clin. Physiol. 17, 401–408 (1997).

102. de Zambotti, M., Nicholas, C. L., Colrain, I. M., Trinder, J. A. & Baker, F. C. Autonomic regulation across phases of the menstrual cycle and sleep stages in women with premenstrual syndrome and healthy controls. Psychoneuroendocrinology 38, 2618–2627 (2013).

103. Pan, Q. et al. Wearable-measured heart rate variability and premenstrual disorder symptoms across menstrual cycle. medRxiv doi:10.1101/2024.10.27.24316196 (2024).

104. Delray, K., Lewis, G. & Hayes, J. F. Tracking mood symptoms across the menstrual cycle in women with depression using ecological momentary assessment and heart rate variability. *BMJ Ment*. Health 28; 10.1136/bmjment-2025-301674 (2025).

105. Van Reen, E. & Kiesner, J. Individual differences in self-reported difficulty sleeping across the menstrual cycle. Arch. Womens Ment. Health 19, 599–608 (2016).

106. Baker, F. C. & Driver, H. S. Self-reported sleep across the menstrual cycle in young, healthy women. J. Psychosom. Res. 56, 239–243 (2004).

107. Baker, F. C. & Driver, H. S. Circadian rhythms, sleep, and the menstrual cycle. Sleep Med. 8, 613–622 (2007).

108. Zheng, H. et al. Actigraphy-defined measures of sleep and movement across the menstrual cycle in midlife menstruating women: Study of women’s health across the nation sleep study. Menopause 22, 66–74 (2015).

109. Li, D. X., Romans, S., De Souza, M. J., Murray, B. & Einstein, G. Actigraphic and self-reported sleep quality in women: associations with ovarian hormones and mood. Sleep Med. 16, 1217–1224 (2015).

110. Costeira, R. et al. Estrogen and COVID-19 symptoms: associations in women from the COVID Symptom Study. PLoS One 16, e0257051; 10.1371/journal.pone.0257051 (2021).

111. Germini, F. et al. Accuracy and acceptability of wrist-wearable activity-tracking devices: systematic review of the literature. J. Med. Internet Res. 24, e30791; 10.2196/30791 (2022).

112. Shin, G. et al. Wearable activity trackers, accuracy, adoption, acceptance and health impact: A systematic literature review. J. Biomed. Inform. 93, 103153; 10.1016/j.jbi.2019.103153 (2019).

113. Fuller, D. et al. Reliability and validity of commercially available wearable devices for measuring steps, energy expenditure, and heart rate: systematic review. JMIR Mhealth Uhealth 8, e18694; 10.2196/18694 (2020).

114. Molnar, G. W. Body temperatures during menopausal hot flashes. J. Appl. Physiol. 38, 499–503 (1975).

115. Freedman, R. R. & Woodward, S. Core body temperature during menopausal hot flushes. Fertil. Steril. 65, 1141–1144 (1996).

116. Kryder, C. How accurate is Oura’s temperature data? Ōura Health Oy https://ouraring.com/blog/temperature-validated-accurate/ (2020).

117. Hasselberg, M. J., McMahon, J. & Parker, K. The validity, reliability, and utility of the iButton® for measurement of body temperature circadian rhythms in sleep/wake research. Sleep Med. 14, 5–11 (2013).

118. Burgess, G. E, 3rd., Cooper, J. R., Marino, R. J. & Peuler, M. J. Continuous monitoring of skin temperature using a liquid-crystal thermometer during anesthesia. South Med. J. 71, 516–518 (1978).

119. Kräuchi, K. & Wirz-Justice, A. Circadian rhythm of heat production, heart rate, and skin and core temperature under unmasking conditions in men. Am. J. Physiol. 267, R819–29 (1994).

120. Driver, H. S., Werth, E., Dijk, D.-J. & Borbély, A. A. The menstrual cycle effects on sleep. Sleep Med. Clin. 3, 1–11 (2008).

121. Miller, D. J., Sargent, C. & Roach, G. D. A validation of six wearable devices for estimating sleep, heart rate and heart rate variability in healthy adults. Sensors (Basel) 22; 10.3390/s22166317 (2022).

122. Kinnunen, H., Rantanen, A., Kenttä, T. & Koskimäki, H. Feasible assessment of recovery and cardiovascular health: accuracy of nocturnal HR and HRV assessed via ring PPG in comparison to medical grade ECG. Physiol. Meas. 41, 04NT01 (2020).

123. Kinnunen, H. O. & Koskimäki, H. The HRV of the ring - comparison of nocturnal HR and HRV between a commercially available wearable ring and ECG. Sleep 41, A120–A120 (2018).

124. Boonya-Ananta, T. et al. Synthetic photoplethysmography (PPG) of the radial artery through parallelized Monte Carlo and its correlation to body mass index (BMI). Sci. Rep. 11, 2570; 10.1038/s41598-021-82124-4 (2021).

125. Osorio-Sanchez, L., May, J. M. & Kyriacou, P. Evaluation of skin pigmentation effect on photoplethysmography signals using a vascular finger phantom with tunable optical and mechanical properties. J. Biomed. Opt. 30, 117002 (2025).

126. Icenhower, A. et al. Investigating the accuracy of Garmin PPG sensors on differing skin types based on the Fitzpatrick scale: cross-sectional comparison study. Front. Digit. Health 7; 10.3389/fdgth.2025.1553565 (2025).

127. Zhu, K. et al. Vision-based heart and respiratory rate monitoring during sleep - a validation study for the population at risk of sleep apnea. IEEE J. Transl. Eng. Health Med. 7, 1900708; 10.1109/JTEHM.2019.2946147 (2019).

128. Hayashi, K., Kawashima, T. & Suzuki, Y. Effect of menstrual cycle phase on the ventilatory response to rising body temperature during exercise. J. Appl. Physiol. (1985) 113, 237–245 (2012).

129. Berryhill, S. et al. Effect of wearables on sleep in healthy individuals: a randomized crossover trial and validation study. J. Clin. Sleep Med. 16, 775–783; 10.5664/jcsm.8356 (2020).

130. Brar, T. K., Singh, K. D. & Kumar, A. Effect of different phases of menstrual cycle on heart rate variability (HRV). J. Clin. Diagn. Res. 9, CC01–4 (2015).

131. Voronova, N. V., Meigal, A. Y., Yelaeva, L. E. & Kuzmina, G. I. Heart rate variability in women during various seasons and phases of the menstrual cycle. Èkol. čeloveka 22, 20–26 (2015).

132. Asgari Mehrabadi, M., et al. Sleep tracking of a commercially available smart ring and smartwatch against medical-grade actigraphy in everyday settings: instrument validation study. JMIR Mhealth Uhealth 8, e20465; 10.2196/20465 (2020).

133. Kubala, A. G. et al. Field-based measurement of sleep: agreement between six commercial activity monitors and a validated accelerometer. Behav. Sleep Med. 18, 637–652 (2020).

134. Nulty, A. K., Chen, E. & Thompson, A. L. The Ava bracelet for collection of fertility and pregnancy data in free-living conditions: An exploratory validity and acceptability study. *Digit*. Health 8; 10.1177/20552076221084461 (2022).

135. Department for Health and Social Care (DHSC). Results of the ‘Women’s Health – Let’s talk about it’ survey. UK Government https://www.gov.uk/government/calls-for-evidence/womens-health-strategy-call-for-evidence/outcome/results-of-the-womens-health-lets-talk-about-it-survey (2022).

136. Kerman, S. Mirror/mirror: women’s reflections on beauty, age and media — authenticity and menopause AARP https://www.aarp.org/pri/topics/aging-experience/older-women-authentic-representation-menopause/ (2025).

137. Hudson, N. The missed disease? Endometriosis as an example of ‘undone science’. Reprod. Biomed. Soc. Online 14, 20–27; 10.1016/j.rbms.2021.07.003 (2022).

138. Meuleman, C. et al. High prevalence of endometriosis in infertile women with normal ovulation and normospermic partners. Fertil. Steril. 92, 68–74 (2009).

139. Nassaralla, C. L. et al. Characteristics of the menstrual cycle after discontinuation of oral contraceptives. J. Womens Health (Larchmt) 20, 169–177 (2011).

140. Blaser, B. L., Weymar, M. & Wendt, J. Alleviating premenstrual symptoms with smartphone-based heart rate variability biofeedback training: a pilot study. Front. Digit. Health 6; 10.3389/fdgth.2024.1337667 (2024).

141. Pasadyn, S. R. et al. Accuracy of commercially available heart rate monitors in athletes: a prospective study. Cardiovasc. Diagn. Ther. 9, 379–385 (2019).

142. Schweizer, T. & Gilgen-Ammann, R. Wrist-worn and arm-worn wearables for monitoring heart rate during sedentary and light-to-vigorous physical activities: device validation study. JMIR Cardio. 9, e67110; 10.2196/67110 (2025).

143. Vidal, C. The ethics of menstrual tracking applications. *Nat*. Hum. Behav. 8, 2076 (2024).

144. Alfawzan, N., Christen, M., Spitale, G. & Biller-Andorno, N. Privacy, data sharing, and data security policies of women’s mHealth apps: scoping review and content analysis. JMIR Mhealth Uhealth 10, e33735; 10.2196/33735 (2022).

145. Dong, Z., Wang, L., Xie, H., Xu, G. & Wang, H. Privacy analysis of period tracking mobile apps in the post-roe v. Wade era. in Proceedings of the 37th IEEE/ACM International Conference on Automated Software Engineering (ASE’22) 1-6 10.1145/3551349.3561343 (Association for Computing Machinery, 2022).

146. Haddaway, N. R., Page, M. J., Pritchard, C. C. & McGuinness, L. A. An R package and Shiny app for producing PRISMA 2020-compliant flow diagrams, with interactivity for optimised digital transparency and Open Synthesis. Campbell Syst. Rev. 18, e1230; 10.1002/cl2.1230 (2022).

147. Namie, T. et al. Menstrual variations of sleep-wake rhythms in healthy women. Sleep Biol. Rhythms 23, 5–12 (2025).

148. Meserve, C. New WHOOP 4.0 Metric: Skin Temperature. WHOOP, Inc https://www.whoop.com/de/en/thelocker/how-whoop-tracks-skin-temperature/ (2021).

149. Gobinath, A. What sensors are on the Ava bracelet? Ava https://www.avawomen.com/avaworld/sensors-ava-bracelet/ (2018).

150. Track your nightly wrist temperature changes with Apple Watch. Apple Inc https://support.apple.com/en-us/102674 (2025).

151. What is the skin temperature feature in Garmin Connect? Garmin Ltd https://support.garmin.com/en-US/?faq=RDXq6E5Iaq9rD1l1kTObf6.

152. How can Fitbit help me track my temperature? Google LLC https://support.google.com/fitbit/answer/14237207?sjid=13360756588868122580-NC.

153. Resting Heart Rate. Ōura Health Oy https://support.ouraring.com/hc/en-us/articles/360025588793-Resting-Heart-Rate.

154. Jasinski, S. R., Rowan, S., Presby, D. M., Claydon, E. A. & Capodilupo, E. R. Wearable-derived maternal heart rate variability as a novel digital biomarker of preterm birth. PLoS One 19, e0295899; 10.1371/journal.pone.0295899 (2024).

155. Haghayegh, S., Khoshnevis, S., Smolensky, M. H. & Diller, K. R. Accuracy of PurePulse photoplethysmography technology of Fitbit Charge 2 for assessment of heart rate during sleep. Chronobiol. Int. 36, 927–933 (2019).

156. Ding, J., Hong, W. & Li, H. Comparison of the accuracy of three wrist-worn tracking devices when measuring heart rate during aerobic exercise. in International Society of Biomechanics in Sports Conference (ISBS ‘22) https://commons.nmu.edu/isbs/vol40/iss1/35 (International Society of Biomechanics in Sports, 2022).

157. Heart rate measurements on my HUAWEI wearable device are inaccurate. HUAWEI https://consumer.huawei.com/en/support/content/en-us00733859/.

158. How accurate is Oura’s respiratory rate? Ōura Health Oy https://ouraring.com/blog/how-accurate-is-ouras-respiratory-rate/?srsltid=AfmBOor3r1JoVjMnBWWGjnoWZGjxjG26xlGPK118jIb0PE2ly8yinlx0 (2020).

159. Capodilupo, E. Understanding respiratory rate: What it is, what’s normal & why you should track it. WHOOP, Inc https://www.whoop.com/qa/en/thelocker/what-is-respiratory-rate-normal/ (2021).

160. Renevey, P., et al. Respiratory and cardiac monitoring at night using a wrist wearable optical system. arXiv 10.1109/EMBC.2018.8512881 (2017).

161. How do I track breathing rate in the Fitbit app? Google LLC https://support.google.com/fitbit/answer/14237113?sjid=13360756588868122580-NC.

162. Natarajan, A. et al. Measurement of respiratory rate using wearable devices and applications to COVID-19 detection. NPJ Digit. Med. 4, 136; 10.1038/s41746-021-00493-6 (2021).

163. Cao, R. et al. Accuracy assessment of Oura Ring nocturnal heart rate and heart rate variability in comparison with electrocardiography in time and frequency domains: comprehensive analysis. J. Med. Internet Res. 24, e27487; 10.2196/27487 (2022).

164. Meisel, L. What your Ava data means. Ava AG https://www.avawomen.com/avaworld/ava-data-means (2017).

165. Grandner, M. A. et al. Performance of a multisensor smart ring to evaluate sleep: in-lab and home-based evaluation of generalized and personalized algorithms. Sleep 46, (2023).

166. What should I know about Fitbit sleep stages? Google LLC https://support.google.com/fitbit/answer/14236712#zippy=%2Chow-does-my-fitbit-device-automatically-detect-my-sleep-stages.

167. Haghayegh, S., Khoshnevis, S., Smolensky, M. H., Diller, K. R. & Castriotta, R. J. Accuracy of wristband Fitbit models in assessing sleep: systematic review and meta-analysis. J. Med. Internet Res. 21, e16273; 10.2196/16273 (2019).

168. HUAWEI Band 8 Principles behind sleep tracking. Huawei https://consumer.huawei.com/en/support/content/en-us15963854/.

169. Mei, L., He, Z. & Hu, L. Accuracy of the Huawei GT2 smartwatch for measuring physical activity and sleep among adults during daily life: instrument validation study. JMIR Form. Res. 8, e59521; 10.2196/59521 (2024).

